# Using Mendelian randomisation to explore the gateway hypothesis: Possible causal effects of smoking initiation and alcohol consumption on substance use outcomes

**DOI:** 10.1101/2021.01.12.21249649

**Authors:** Zoe E. Reed, Robyn E. Wootton, Marcus R. Munafò

## Abstract

**Background and Aims:** Initial use of drugs such as tobacco and alcohol may lead to subsequent more problematic drug use – the ‘gateway’ hypothesis. However, observed associations may be due to a shared underlying risk factor, such as trait impulsivity. We used bidirectional Mendelian Randomisation (MR) to test the gateway hypothesis.

**Design:** Our main method was inverse-variance weighted (IVW) MR, with other methods included as sensitivity analyses (where consistent results across methods would raise confidence in our primary results). MR is a genetic instrumental variable approach used to support stronger causal inference in observational studies.

**Setting:** European ancestry individuals.

**Participants:** Genome-wide association summary data for smoking initiation, alcoholic drinks per week, cannabis use and dependence, cocaine and opioid dependence (N=1,749 to 1,232,091).

**Measurements:** Genetic variants for exposure.

**Findings:** We found evidence of causal effects from smoking initiation to increased drinks per week (IVW: β=0.06; 95% CI=0.03 to 0.09; p=9.44×10^−06^), cannabis use (IVW: OR=1.34; 95% CI=1.24 to 1.44; p=1.95×10^−14^), and cannabis dependence (IVW: OR=1.68; 95% CI=1.12 to 2.51; p=0.01). We also found evidence of an effect of cannabis use on increased likelihood of smoking initiation (IVW: OR=1.39; 95% CI=1.08 to 1.80; p=0.01). We did not find evidence of an effect of drinks per week on other substance use outcomes, except weak evidence of an effect on cannabis use. We found weak evidence of an effect of opioid dependence on increased drinks per week (IVW: β=0.002; 95% CI=0.0005 to 0.003; p=8.61×10^−03^).

**Conclusions:** Smoking initiation may lead to increased alcohol consumption, cannabis use and dependence. Cannabis use may also lead to smoking initiation, and opioid dependence to alcohol consumption. However, given tobacco and alcohol use typically begin before other drug use, these results may reflect a shared risk factor, or a bidirectional effect for cannabis use. Further research should explore potentially shared risk factors.

## Introduction

Illicit substance use and substance use disorders result in a substantial global burden on a range of health conditions (1,2). Identifying causal risk factors in the development of problematic substance use is important for designing successful interventions and preventing subsequent health problems.

The gateway hypothesis, in its simplest form, is the theory that initial use of legal ‘gateway’ drugs, including tobacco and alcohol, may lead to illicit drug use, such as cannabis, cocaine and opioids (3–5). Previous studies have found associations between smoking initiation and use of alcohol (6), cannabis (7,8), cocaine (9) and opioids (10). Studies also suggest alcohol as a possible gateway drug (11–14). Given that tobacco and alcohol consumption are likely to both occur initially during adolescence, and typically before other drug taking, it is important to investigate both as potential gateway drugs. Prospective studies also support the gateway hypothesis for these outcomes (7,8,15–17), suggesting possible causal relationships. Substance use behaviours are moderately heritable (21% to 72% in twin studies) (18–22). Genetic correlations have also been found between different substance use phenotypes (*r*_*G*_ 0.35 to 0.66) (23–26).

Whilst these studies may support the gateway hypothesis, it is equally plausible that there are underlying shared risk factors, for example risk taking or impulsive behaviours. Previous studies have reported an association of ADHD with substance use outcomes (27,28), and ADHD genetic risk with smoking initiation (29,30), supporting impulsivity as a potential shared risk factor, although others - such as risk taking, or adverse childhood experiences - could also lead to these outcomes. In terms of establishing whether the relationships between smoking and alcohol and other substance use are causal, there is some evidence (e.g., from randomised controlled trials) that smoking cessation may result in reduced substance use or abstinence (31); supporting a possible causal effect of smoking on substance use outcomes.

Mendelian Randomisation (MR) is a well-established method for causal inference, based on instrumental variable (IV) analysis, which attempts to overcome issues of residual confounding and reverse causation (32–35). MR uses genetic variants, assigned randomly at conception, as IVs for an exposure to estimate the causal relationship with an outcome. In two-sample MR (36) the single nucleotide polymorphism (SNP)-exposure and SNP-outcome estimates are obtained from independent sample genome-wide association studies (GWAS) to estimate possible causal effects. Previous MR studies examining this relationship looked at cannabis use only, and used smaller GWAS sample sizes than in the current study. One study found weak evidence of a causal effect of smoking initiation on cannabis use (37), whilst the other found no evidence (38). Incorporating larger GWAS and a range of substance use outcomes may improve power to detect causal effects and provide better evidence as to whether these relationships are due to a gateway effect.

We applied this two-sample MR approach to investigate the possible causal effect between both smoking initiation and alcohol consumption (defined as drinks per week) and substance use outcomes of cannabis use and dependence, cocaine dependence and opioid dependence. We refer to these outcomes as illicit substance use, although we acknowledge that cannabis is not illegal in all jurisdictions. We also examined the association between smoking initiation and alcohol consumption. We used a bidirectional approach (Figure 1) to assess whether there is evidence supporting the gateway hypothesis (i.e., that smoking initiation/alcohol consumption can lead to use of other substances and dependence), or whether there is evidence of a shared risk factor. Some pathways (e.g., from opioid use to smoking initiation) are unlikely, so analyses in this direction acted more as a sensitivity analysis, which could help identify a shared risk factor rather than a causal effect.

**Figure 1.**
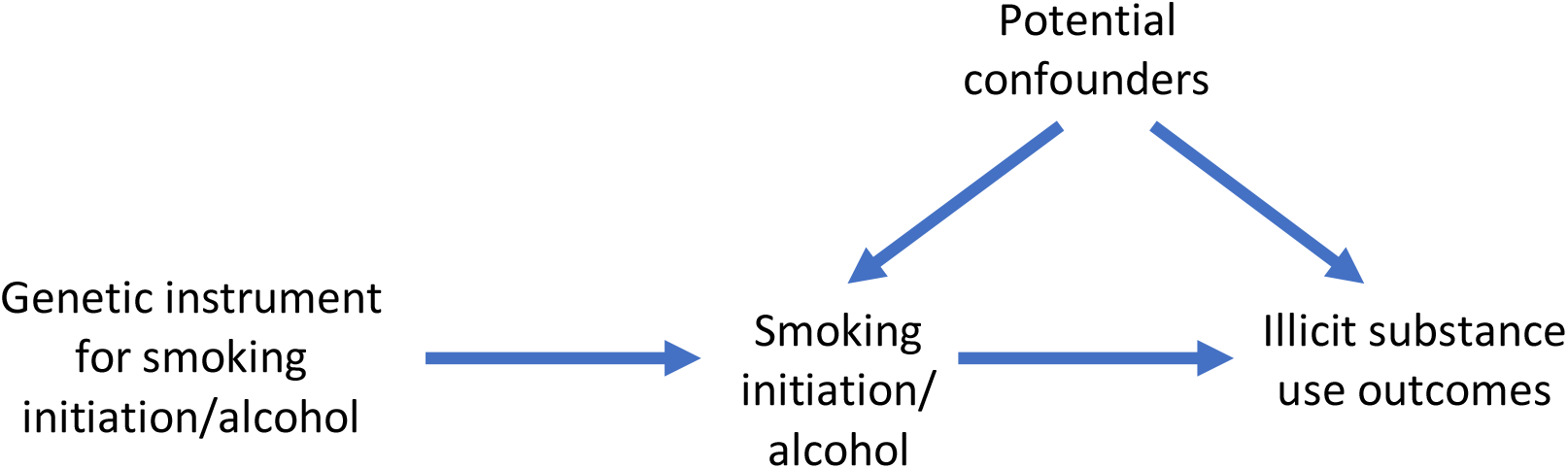
Bidirectional two-sample Mendelian randomisation between smoking initiation/alcohol consumption and illicit substance use outcomes. A directed acyclic graph (DAG) for the causal effect between smoking initiation/alcohol consumption and illicit substance use outcomes. Evidence of a causal effect in the other direction may indicate a bidirectional effect, or a common underlying risk factor.

## Methods

### Data sources

We used GWAS summary statistics obtained from several consortia and other samples, the details of which are shown in Table 1, along with the variance explained by genome-wide significant SNPs and SNP heritabilities where these were reported. GWAS were conducted in samples of European ancestry. Sample overlap should be avoided or reduced, so as not to bias the estimates towards a more conservative effect estimate (39). Therefore, we used GWAS with certain samples excluded from the consortia (see Table 1).

**Table 1.** GWAS used for two sample Mendelian Randomisation.

| Phenotype | Reference | Consortium or sample | Excluded samples | Final sample size | R<sup>2</sup> and SNP heritability<sup>1</sup> |
| --- | --- | --- | --- | --- | --- |
| **Smoking initiation (ever/never)** | Liu et al, 2019 (23) | GSCAN (23andMe, ALSPAC, ARIC, BEAGESS, BLTS, CADD, COGEND, COPDGene, deCODE, EGCUT, FHS, FinnTwin, GERA, GfG, Harvard, HRS, HUNT, MCTFR, MESA, METSIM, NESCOG, NAG-FIN, NTR, QIMR, SardinIA, UK Biobank, WHI) | Dependent on analysis, published GWAS summary statistics were used other than for analyses with outcomes for i) cannabis use where only the 23andMe sample was included and ii) drinks per week where 23andMe was excluded. | All samples = 1,232,091<br>i) cannabis use = 599,289<br>ii) drinks per week = 632,802 | R<sup>2</sup>=2.32%<br>h<sup>2</sup>=7.8% |
| **Drinks per week** | Liu et al, 2019 (23) | GSCAN | Dependent on analysis, published GWAS summary statistics were used other than for analyses with outcomes for i) cannabis use where only the 23andMe sample was included. | All samples= 941,280<br>i) cannabis use = 403,931 | R<sup>2</sup>=0.19%<br>h<sup>2</sup>=4.2% |
| **Cannabis use (ever/never)** | Pasman et al, 2018 (41) | ICC (ALSPAC, BLTS, CADD, EGCUT1, EGCUT2, FinnTwin, HUVH, MCTFR, NTR, QIMR, TRAILS, Utrecht, Yale Penn EA), UK Biobank and 23andMe | 23andMe | 162,082 | R<sup>2</sup>=0.15%<br>h<sup>2</sup>=11% |
| **Cannabis dependence** | Agrawal et al, 2017 (42) | CATS, COGA-cc, COGA-f, OZALC, SAGE | NA | 2,080 cases and 6,435 exposed controls | Not reported |
| **Cocaine dependence (DSM-IV criteria)** | Gelernter et al, 2014 (43) | Yale (APT Foundation), University of CT, MUSC, McLean Hospital, University of Pennsylvania | NA | 1,809 cases and 292 exposed controls | Not reported |
| **Opioid dependence (DSM-IV criteria)** | Gelernter et al, 2014 (44) | Yale (APT Foundation), University of CT, MUSC, McLean Hospital, University of Pennsylvania | NA | 1,383 cases and 366 exposed controls | Not reported |

<sup>1</sup>The $r^2$ reported was for genome-wide significant SNPs only and the SNP heritability ( $h^2$ ) was for all SNPs.
GSCAN= GWAS & Sequencing Consortium of Alcohol and Nicotine use, ALSPAC=Avon Longitudinal Study of Parents and Children, ARIC=Atherosclerosis Risk in Communities, BEAGESS=The Barrett's and Esophageal Adenocarcinoma Genetic Susceptibility Study, BLTS=Brisbane Longitudinal Twin Study, CADD=Center on Antisocial Drug Dependence, COGEND=Collaborative Genetic Study of Nicotine Dependence, COPDGene=Genetics of Chronic Obstructive Pulmonary Disease, EGCUT=Estonian Genome Center, FHS=Framingham Heart Study, FinnTwin and NAG-FIN =Finnish Twin Cohort, GERA=Genetic Epidemiology Research in Adult Health and Aging, GfG=Genes for Good, Harvard, HRS=Health and Retirement Study, HUNT=The Nord-Trøndelag Health Study, MCTFR=Minnesota Center for Twin and Family Research, MESA=Multi-Ethnic Study of Atherosclerosis, METSIM=Metabolic Syndrome in Men, NESCOG=Netherlands Study on Cognition, Environment and Genes, NTR=Netherlands Twin Register, WHI=Women's Health Initiative, COGA= Collaborative Study on the Genetics of Alcoholism, SAGE= Study of Addictions: Genes and Environment, OZALC= Australian Alcohol, Nicotine Addiction Genetics and Childhood Trauma, CATS= Comorbidity and Trauma Study.

#### Smoking initiation

The smoking initiation GWAS (23) identified 378 conditionally independent genome-wide significant SNPs associated with ever being a smoker i.e., where participants reported ever being a regular smoker in their life. See Supplementary Materials for further details. The total sample size was 1,232,091 for the GSCAN consortium, however the sample size for the GWAS in each of our analyses varied to try and avoid sample overlap (see Table 1). Full genome-wide summary statistics were only publicly available without 23andMe. We requested 23andMe summary statistics separately and meta-analysed them with the publicly available data to recreate the original full GWAS summary statistics. The meta-analysis was conducted using the genome-wide association meta-analysis (GWAMA) software (40).

#### Drinks per week

The drinks per week GWAS (23) identified 99 independent genome-wide significant SNPs associated with the average number of alcoholic drinks consumed per week. See Supplementary Materials for further details.

#### Cannabis use

The cannabis use GWAS (41) identified 8 independent genome-wide significant SNPs associated with ever using cannabis. See Supplementary Materials for further details.

#### Cannabis dependence

The cannabis dependence GWAS (42) did not identify any genome-wide significant SNPs associated with cannabis dependence. Cases were established based on meeting three or more criteria for Diagnostic and Statistical Manual of Mental Disorders, 4th edition (DSM-IV) cannabis dependence.

#### Cocaine dependence

The cocaine dependence GWAS (43) identified one genome-wide significant SNP associated with cocaine dependence. All participants were interviewed using the Semi-structured Assessment for Drug Dependence and Alcoholism (SSADA) and cocaine dependent cases were established based on responses according to the DSM-IV criteria and reflect lifetime cocaine dependence.

#### Opioid dependence

The opioid dependence GWAS (44) did not identify any genome-wide significant SNPs associated with opioid dependence. All participants were interviewed using the SSADA and opioid dependent cases were established based on responses according to the DSM-IV criteria and reflect lifetime opioid dependence.

Units for all binary measures were in log odds ratios and for the continuous drinks per week measure were per SD increase in the number of drinks per week.

### Statistical analyses

MR was used to assess whether relationships may be causal by using genetic variants as IV proxies for the exposures. Further details can be found in the supplementary materials. Two-sample MR was conducted in R (version 4.0.0) (45) using the TwoSampleMR package (version 0.5.3) (46,47). Genome-wide significant SNPs were selected as instruments for the smoking initiation, alcohol and cannabis use exposures. However, where cocaine, opioid and cannabis dependence were the exposures there were either too few or no genome-wide significant SNPs, so we used a less stringent threshold of 1×10^−05^.

Multiple MR methods were used to assess the causal effects of: i) the exposure of smoking initiation/alcohol consumption on illicit substance use outcomes, and ii) illicit substance use exposures on smoking initiation/alcohol consumption. These were inverse-variance weighted (IVW) (48), MR-Egger (49), weighted median (50), simple mode and weighted mode (51) MR methods. We were interested in the question of whether there is evidence of causal effects. We were concerned with the strength of evidence for an effect, as opposed to the effect estimate and considered whether the direction of effect was as predicted and the strength of statistical evidence against the null. To do this we interpreted the p-value as a continuous measure of statistical evidence (52) and considered whether our results were consistent across different MR approaches. The IVW approach was our main method, with the others being sensitivity analyses which make different assumptions. We describe our findings in terms of lack of evidence, weak evidence, evidence, or strong evidence of an effect, accounting for all these factors. The sensitivity methods have less statistical power than the IVW approach; therefore we considered all results, and the consistency of the direction of effect observed across analyses. Inconsistent results for these sensitivity analyses may indicate that some MR assumptions are violated (e.g., pleiotropic pathways are operating). Specifically, the IVW method constrains the intercept to be zero and assumes all SNPs are valid instruments with no horizontal pleiotropy. Horizontal pleiotropy can be problematic as MR assumptions may be violated if the SNPs affect the outcome via a different pathway. Therefore, we included additional tests which can detect whether horizontal pleiotropy may be present. For example, we included results for the Cochran’s test of heterogeneity, which assesses whether there is heterogeneity in the SNP_exposure_ – SNP_outcome_ associations for each SNP included in the instrument. If there is evidence of heterogeneity this may indicate possible horizontal pleiotropy.

The MR-Egger method tests whether there is overall directional pleiotropy by not constraining the intercept, where a non-zero intercept indicates directional horizontal pleiotropy. We also used the Rucker’s Q-test to assess heterogeneity in the MR-Egger estimates for individual SNPs, similar to the Cochran’s test. The weighted median method provides an estimate under the assumption that at least 50% of the SNPs are valid instruments (i.e., satisfy the IV assumptions). Finally, the mode-based approaches provide an estimate for the largest cluster of similar SNPs, where the SNPs not in that cluster could be invalid, with the weighted method taking into account the largest weights of SNPs.

Additionally, we also estimated effects for single SNP and leave-one-out analyses and plotted these results, where there was evidence for a causal effect.

We also estimated the mean F-statistic, unweighted and weighted I-squared values for each of the analyses (53). The F-statistic represents instrument strength, where a value under 10 may indicate a weak instrument (53). The I-squared value falls between zero and one and indicates the amount of bias in the ‘NO Measurement Error’ (NOME) assumption in the MR-Egger estimate. If bias was apparent, we ran run simulation extrapolation (SIMEX) corrections and present these in place of the MR-Egger results, or if the bias was too large neither were presented (see supplementary materials for further details).

Finally, we conducted multivariable MR (MVMR) to investigate whether the causal effect of smoking initiation was independent of that for the drinks per week exposure for any illicit substance use outcomes where both exposures were associated with the outcome. MVMR is an extension of MR that estimates the causal effect of multiple exposures on an outcome and assesses whether each exposure is independent of the others (54). Please note that our analyses were not pre-registered and therefore our results should be considered exploratory.

## Results

### Evidence of causal effects of smoking initiation on illicit substance use outcomes

Our two-sample MR results (Table S2 and Figure 2) indicated there was evidence for a causal effect of smoking initiation on increased drinks per week (IVW: β=0.06; 95% CI 0.03 to 0.09; p-value=9.44×10^−06^). The I-squared values (Table S1) suggest the MR Egger method was unsuitable; therefore, results are not presented for this. Results were in a consistent direction with evidence of a causal effect across the different MR analyses (see also Figure S1). We observed evidence of heterogeneity in results for the IVW method (see also Figure S2), but this was not necessarily indicative of horizontal pleiotropy (see also Figure S3). Leave-one-out analyses did not reveal that any single SNP was driving the association (Figure S4).

**Figure 2.**
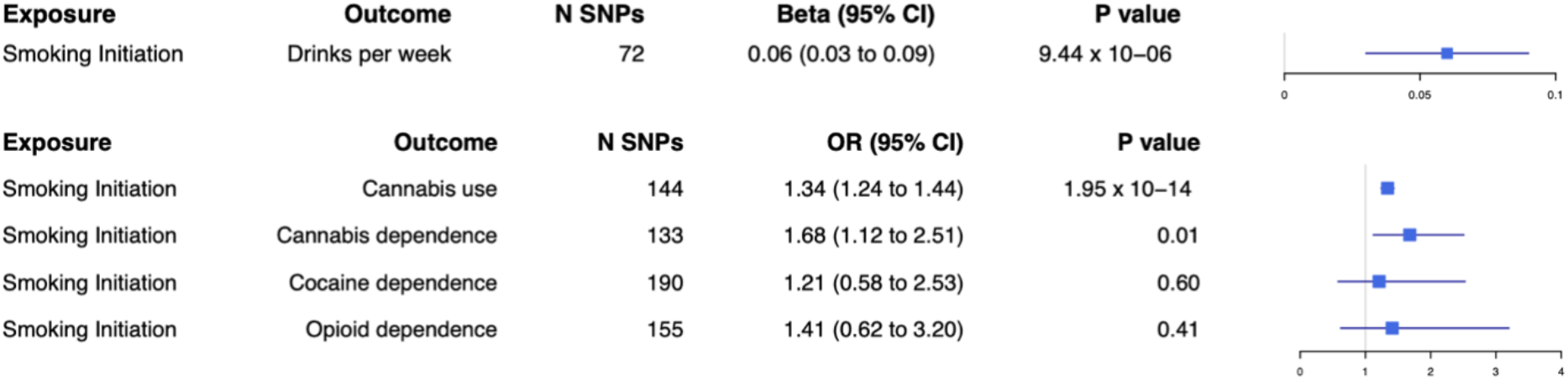
Forest plot for two-sample Mendelian randomisation with smoking initiation as the exposure. Causal effects from the inverse-variance weighted Mendelian randomisation method where smoking initiation is the exposure. Effect estimates are presented as beta or odds ratios (OR) depending on whether the outcome was continuous or binary, with 95% confidence intervals (CI). SNP=single nucleotide polymorphism.

We also found evidence of a causal effect of smoking initiation on cannabis use (IVW: OR=1.34; 95% CI=1.24 to 1.44; p-value=1.95×10^−14^). Results were in a consistent direction across MR analyses (see also Figure S5), although evidence for this was only found additionally for the weighted median method. There was evidence of heterogeneity with both the IVW and MR Egger methods (see also Figure S6) but not horizontal pleiotropy (see also Figure S7). Leave-one-out analyses did not reveal that any single SNP was driving the association (Figure S8).

We found evidence of a causal effect of smoking initiation on cannabis dependence (IVW: OR=1.68; 95% CI 1.12 to 2.51; p-value=0.01). Results were in a consistent direction for the SIMEX adjusted MR Egger and weighted median methods (Table S1), although evidence for these was weak (see also Figure S9). There was no evidence of heterogeneity or horizontal pleiotropy (also see Figures S10 and S11). Leave-one-out analyses did not reveal that any single SNP was driving the association (Figure S12).

Finally, we did not find evidence of a causal effect of smoking initiation on cocaine dependence (IVW: OR=1.21; 95% CI 0.58 to 2.53; p-value=0.60) or opioid dependence (IVW: OR=1.41; 95% CI 0.62 to 3.20; p-value=0.41) with any of the MR analyses, except for weak evidence for the SIMEX adjusted (Table S1) MR-Egger for cocaine dependence. There was no evidence of heterogeneity or horizontal pleiotropy for cocaine or opioid dependence.

### Causal effects of illicit substance use exposures on smoking initiation

For the direction of illicit substance use to smoking initiation (Table S3 and Figure 3) we found evidence of a causal effect of cannabis use on smoking initiation (IVW: OR=1.39; 95% CI=1.08 to 1.80; p-value=0.01) for all MR analyses except MR Egger. Results were in a consistent direction across MR analyses (see also Figure S13). We observed evidence of heterogeneity in these results for the IVW and MR Egger methods (see also Figure S14), but not horizontal pleiotropy (see also Figure S15). Leave-one-out analyses did not reveal that any single SNP was driving the association (Figure S16).

**Figure 3.**
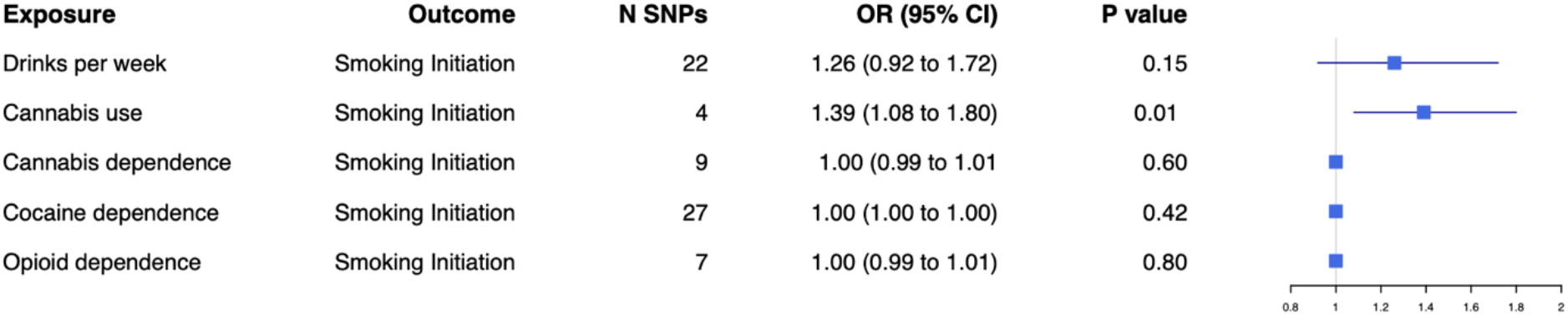
Forest plot for two-sample Mendelian randomisation with smoking initiation as the outcome. Causal effects from the inverse-variance weighted Mendelian randomisation method where smoking initiation is the outcome. Effect estimates are presented as odds ratios (OR) with 95% confidence intervals (CI). SNP=single nucleotide polymorphism.

We did not find any evidence of a causal effect of drinks per week, (IVW: OR=1.26; 95% CI=0.92 to 1.72; p-value=0.15), cannabis dependence (IVW: OR=1.00; 95% CI=0.99 to 1.01; p-value=0.60), cocaine dependence (IVW: OR=1.00; 95% CI=1.00 to 1.00; p-value=0.42) or opioid dependence (IVW: OR=1.00; 95% CI=0.99 to 1.01; p-value=0.80) on smoking initiation, for any of the MR analyses.

### Causal effects of drinks per week on illicit substance use outcomes

When examining whether there was evidence for causal effects of alcohol consumption (drinks per week) on the illicit substance use phenotypes (Table S4 and Figure 4) we did not find any evidence for the IVW approach for cannabis use (IVW: OR=0.55; 95% CI=0.16 to 1.93; p-value=0.35), although there was some evidence of a causal effect with the other MR analyses. We did not find evidence of a causal effect on cannabis dependence (IVW: OR=2.73; 95% CI=0.62 to 11.95; p-value=0.18), cocaine dependence (IVW: OR=0.50; 95% CI=0.09 to 2.79; p-value=0.43) or opioid dependence (IVW: OR=0.38; 95% CI=0.06 to 2.41; p-value=0.30).

**Figure 4.**
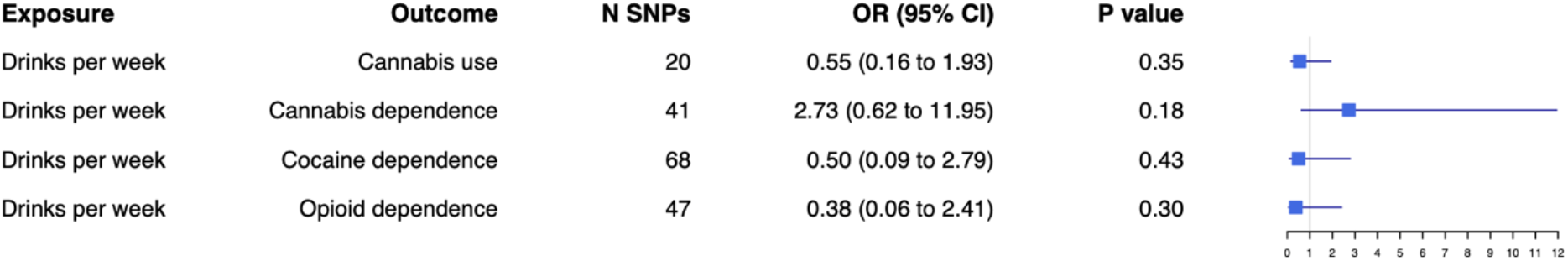
Forest plot for two-sample Mendelian randomisation with drinks per week as the exposure. Causal effects from the inverse-variance weighted Mendelian randomisation method where drinks per week is the exposure. Effect estimates are presented as odds ratios (OR) with 95% confidence intervals (CI). SNP=single nucleotide polymorphism.

### Causal effects of illicit substance use exposures on drinks per week

For the reverse direction (Table S5 and Figure 5) we did not find evidence of a causal effect of cannabis use (IVW: β=0.03; 95% CI=-0.009 to 0.07; p-value=0.14), cannabis dependence (IVW: β=-0.0003; 95% CI=-0.003 to 0.002; p-value=0.80) or cocaine dependence (IVW: β=0.0007; 95% CI=-0.00007 to 0.001; p-value=0.08) on drinks per week.

**Figure 5.**
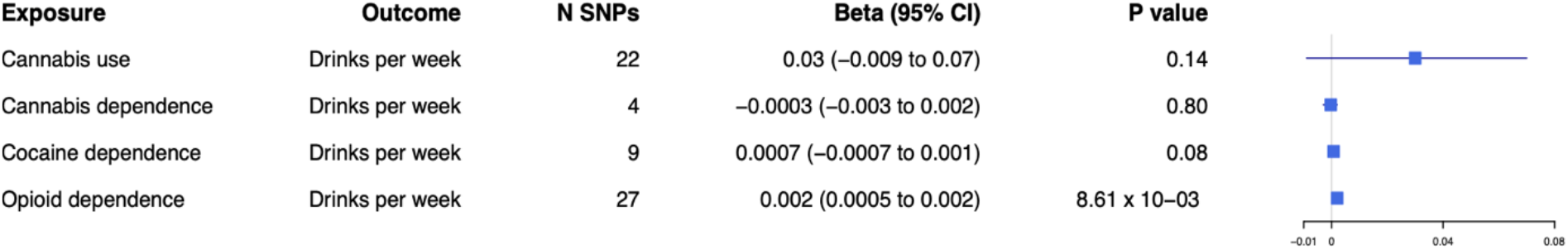
Forest plot for two-sample Mendelian randomisation with drinks per week as the outcome. Causal effects from the inverse-variance weighted Mendelian randomisation method where drinks per week is the outcome. Effect estimates are presented as beta with 95% confidence intervals (CI). SNP=single nucleotide polymorphism.

There was weak evidence to suggest a causal effect of opioid dependence on drinks per week (IVW: β=0.002; 95% CI=0.0005 to 0.003; p-value=8.61×10^−03^), although the effect size was very small, and this was not found for any other MR analyses (see also Figure S17). There was no evidence of heterogeneity (see also Figure S18) or horizontal pleiotropy (Figure S19). Leave-one-out analyses did not reveal that any single SNP was driving the association (Figure S20).

### Multivariable Mendelian Randomisation analysis for cannabis use

We conducted MVMR analysis for cannabis use only due to evidence of a causal effect of smoking initiation on cannabis use and weak evidence of a causal effect of drinks per week on cannabis use. We found evidence of a direct effect of smoking initiation, independent of drinks per week on cannabis use (OR=1.35; 95% CI=1.25 to 1.46; p-value=3.67×10^−12^). This result was similar to that from the two-sample MR model. However, there was no evidence of a direct effect of drinks per week on cannabis use (OR=0.71; 95% CI=0.29 to 1.76; p-value=0.47).

## Discussion

We examined whether there was evidence for causal effects of smoking initiation and alcohol consumption on cannabis use and dependence on cannabis, cocaine and opioids, which may support the ‘gateway’ hypothesis. We also examined the reverse direction, where evidence of an association, particularly in both directions may be indicative of an underlying common risk factor.

Our main findings were those for cannabis use and dependence, which suggest that ever smoking may act as a gateway to subsequent cannabis use and perhaps even dependence, although evidence was weaker for the latter. This supports previous observational studies demonstrating an association between these phenotypes (7,8,55) and is in line with previous findings suggesting tobacco is a gateway drug to other more problematic substance use (5,6,8,10). Our MR analyses support stronger causal inference, although further triangulation with other study designs would strengthen this. Previous literature also suggests that alcohol consumption may be causally associated with cannabis use; however, our MVMR results suggest no evidence for independent effects of alcohol consumption, only evidence for a causal effect of smoking initiation on cannabis use.

We also found evidence for a potential causal pathway from cannabis use to smoking initiation. It has been previously suggested that cannabis use may act as gateway to tobacco use, possibly due to the form in which cannabis is used, i.e., if smoked with tobacco (56). However, our finding of potential causal pathways between cannabis use and smoking initiation in both directions may suggest this association is due to an underlying common risk factor, as opposed to either being a gateway drug. We found that all the SNPs used in the cannabis use instrument, except one, are in LD with genome-wide significant SNPs in the smoking initiation GWAS (r^2^>0.27, 250kb window for 3 SNPs). As this the genetic instruments may be overlapping, this does not help us disentangle the reason behind this relationship.

There are several potential reasons for our results: 1) a causal effect of smoking initiation on cannabis use, 2) a causal effect of cannabis use on smoking initiation, 3) a bidirectional effect, 4) an underlying shared risk factor, 5) horizontal pleiotropy (although our sensitivity analyses suggested this was not biasing results), and 6) confounding due to LD. Without further understanding of the biological function of these genetic variants it is difficult to conclude which of these explanations (which are not mutually exclusive) could be true here and this has been discussed previously in relation to mental health behavioural risk factors (57,58). Previous studies have suggested that impulsive or risk-taking behaviours may be associated with smoking initiation and substance use (59–61). Additionally, cannabis use may capture underlying risk-taking behaviours more than the dependence measures and this may be why we see a more consistent association with this measure. Further research is needed to establish whether there could be an underlying common cause, and if this might be related to risk-taking behaviours. Other potential shared risk factors should also be considered, and these may be genetic or environmental in origin and may vary between different illicit substance use phenotypes. In addition, it may be the case that smoking initiation, for example, only acts as a gateway to other substances in the presence of mediators such as stressful life events or adverse circumstances. Therefore, the mechanisms behind these associations need to be examined further, and the possibility of a bidirectional relationship should also be considered.

We found a potential causal effect of smoking initiation on increased drinks per week, but did not find an association in the reverse direction. It is plausible that an underlying risk-taking behaviour may affect alcohol consumption via smoking. However, a biological mechanism behind this association should also be considered and studied further. Finally, we did see weak evidence of a causal effect of opioid dependence on increased drinks per week; however, due to the low power for the opioid dependence GWAS and the small effect size we would interpret this with caution. Opioid dependence (compared with ever use) is less likely explained by underlying risk-taking behaviour. Therefore, research into alternative shared risk factors is warranted. It may be the case that opioid dependence does have a causal effect on increased alcohol use, and this also warrants further investigation.

### Limitations

Our study is the first, to our knowledge, to examine whether causal pathways may exist between smoking initiation/alcohol consumption and various illicit substance use phenotypes, using an MR approach. However, there are several limitations to note, for example some of our analyses may be limited in their power to detect a causal effect. This is particularly the case where the dependence measures were the exposures, as the GWAS discovery samples were much smaller than those for drinks per week, cannabis use and smoking initiation. Additionally, we used a less stringent p-value threshold of 1×10^−05^ due to a low number of genome-wide significant SNPs. Therefore, these instruments may be less robustly associated with the exposure, and pleiotropy could be introduced. For our results with the dependence variables as exposures, the CIs were very narrow which could be a result of the relaxed p-value thresholds used. However, the absence of evidence here does not mean we can exclude the possibility of an effect for this relationship. Furthermore, the lower number of SNPs used for the dependence exposures may mean the instruments are weak, which may be particularly problematic for MR-Egger. For our finding of an effect of opioid dependence on drinks per week, additional caution should be taken when interpreting this result as the opioid dependence exposure is a dichotomised variable for an underlying latent risk factor. Thus, the estimate here is less interpretable than for our other results and instead focus should be on the direction and evidence of an effect as opposed to the effect size. Therefore, our dependence results should be interpreted with caution revisited once larger GWAS become available.

We also found some evidence of heterogeneity and horizontal pleiotropy for different analyses meaning that these results should be interpreted in light of this, as some of the SNPs used may be associated with the outcome other than via the exposure. However, the additional MR analyses, which account for this, were generally in the same direction as our main results, although we were unable to formally test for directional pleiotropy in some cases where the I-squared estimate was low. In cases where the IVW shows evidence for a causal effect, but results are inconsistent across the sensitivity analyses, this may be indicative of pleiotropy. However, inconsistent effects across sensitivity analyses and no evidence from the IVW is more likely to reflect a lack of evidence for an effect.

Another consideration is that the MR instruments used may not be valid for smoking as they may be picking up risk-taking behaviours more than smoking itself (62). Therefore, it would be useful to examine this further with other smoking related phenotypes such as smoking heaviness. Additionally, whilst we tried to avoid sample overlap, there was still some for the cannabis use GWAS (17% of the sample was also present in the GWAS for smoking initiation and drinks per week). Sample overlap could bias estimates towards a more conservative effect estimate (39), which should be considered when interpreting our results.

The MR analysis itself is subject to several limitations (33). For example, the GWAS used for MR may suffer from ‘Winner’s curse’, where the SNP-exposure estimates may be overestimated, due to selecting SNPs with the smallest p-values and biasing the MR estimate towards the null. Thus, interpreting the direction of effect as opposed to the effect size itself is more valid here. The effect estimate may also be biased by trait heterogeneity, for example, different aspects of substance use behaviours may be associated with the same genetic variants and therefore it is difficult to get a precise estimate for a single aspect of any substance use behaviour.

Finally, our results should be considered in the context of the multiple potential causal pathways that we have investigated.

### Conclusion

Whilst our findings support the gateway hypothesis to some extent, they also point to a potential underlying common risk factor and with better powered GWAS or those with more precise instruments and additional research we may be able to interrogate this further. Triangulating our results with other approaches would help answer this question (63,64). By doing so we may be able to identify risk factors to substance use which could ultimately help with intervention design.

## Data Availability

The data used in this study are publicly available GWAS data which can be obtained from relevant data repositories or directly from the authors. Access for data from 23andMe needs to be requested and can be done here: https://research.23andme.com/dataset-access/. GWAS data for smoking initiation and drinks per week not including 23andMe can be found here: https://conservancy.umn.edu/handle/11299/201564. GWAS data for cannabis use can be obtained here: https://www.ru.nl/bsi/research/group-pages/substance-use-addiction-food-saf/vm-saf/genetics/international-cannabis-consortium-icc/. GWAS data for cannabis dependence, cocaine dependence and opioid dependence were obtained directly from the authors.

## Acknowledgements

We thank all the contributors to the consortia we have used GWAS results from in our analyses. We would like to thank the research participants and employees of 23andMe for making this work possible.

