## Supplementary materials for "Using Mendelian randomisation to explore the gateway hypothesis: Possible causal effects of smoking initiation and alcohol consumption on substance use outcomes"

Supplementary methods

*Smoking initiation*

The smoking initiation phenotype was measured in several different ways, and included questions about whether the participant had smoked over 100 cigarettes in their life, whether they had smoked every day for at least one month and whether they smoked regularly. The binary variable for ever smoking was constructed based on these variables.

*Drinks per week*

This measure for the average number of alcoholic drinks consumed per week did not distinguish between types of alcohol and for any study that used a range in their questions the midpoint was used. Participants were asked questions relating to the number of alcoholic drinks consumed in the past week and on average per week in the past year. The resulting data was log transformed prior to the GWAS.

*Cannabis use*

For the International Cannabis Consortium (ICC) study, the data relating to cannabis use were obtained from self-report questionnaires and recoded as ever versus never for cannabis use in their lifetime. Similarly, UK Biobank participants were asked whether they had used cannabis in an online follow-up study.

*Mendelian Randomisation*

Mendelian Randomisation (MR) is a causal inference method which uses genetic variants as instrumental variables (IVs) for an exposure of interest to estimate the causal relationship with an outcome of interest. For MR to be valid, several assumptions must be met; that the IV and the exposure are associated, the IV is only associated with the outcome via the exposure and that the IV is not associated with potential confounders of the exposure-outcome association.

In our two-sample MR analyses, the SNPs we used as genetic instruments were clumped to ensure independence for linkage disequilibrium (LD) using an r^2^ of 0.001 and a window of 10000 kb. The GWAS summary statistics for these SNPs were harmonised so that the effect of the SNPs were relative to the same allele.

*I-squared values*

A value of 0.9 or above would indicate that there is minimal bias in the MR-Egger estimate. For values between 0.6 and 0.9 we have run simulation extrapolation (SIMEX) corrections and present these in place of the MR-Egger results. Specifically, we present these for the I-squared with the highest value out of the weighted and unweighted I-squared. SIMEX allows for the calculation of bias-adjusted point estimates for MR-Egger. However, for any values below 0.6 (where this bias is too large) it is not appropriate to run the SIMEX correction. We have indicated where this is the case, as these MR Egger results cannot be interpreted with confidence.

Table S1. Mean F-statistic, weighted and unweighted I-squared results for all of the MR analyses.

| **Exposure** | **Outcome** | **Mean F-statistic** | **Unweighted I-squared** | **Weighted I-squared** |
| --- | --- | --- | --- | --- |
| Smoking initiation | Drinks per week | 32.75 | 0.33 | 0.14 |
| Smoking initiation | Cannabis use | 47.64 | 0.82 | 0.95 |
| Smoking initiation | Cannabis dependence | 51.03 | 0.7 | 0.65 |
| Smoking initiation | Cocaine dependence | 49.28 | 0.67 | 0.62 |
| Smoking initiation | Opioid dependence | 49.75 | 0.67 | 0.61 |
| Drinks per week | Smoking initiation | 81.74 | 0.97 | 0.98 |
| Cannabis use | Smoking initiation | 40.76 | 0.09 | 0 |
| Cannabis dependence | Smoking initiation | 22.7 | 0.45 | 0 |
| Cocaine dependence | Smoking initiation | 22.50 | 0.72 | 0.57 |
| Opioid dependence | Smoking initiation | 23.04 | 0 | 0 |
| Drinks per week | Cannabis use | 84.42 | 0.97 | 0.98 |
| Drinks per week | Cannabis dependence | 43.07 | 0.73 | 0.52 |
| Drinks per week | Cocaine dependence | 69.71 | 0.96 | 0.95 |
| Drinks per week | Opioid dependence | 72.5 | 0.97 | 0.96 |
| Cannabis use | Drinks per week | 40.76 | 0.09 | 0 |
| Cannabis dependence | Drinks per week | 22.7 | 0.45 | 0 |
| Cocaine dependence | Drinks per week | 22.50 | 0.72 | 0.38 |
| Opioid dependence | Drinks per week | 23.04 | 0 | 0 |

*The F-statistic is indicative of instrument strength, where a value under 10 may indicate a weak instrument. The I squared indicates the amount of bias in the ‘NO Measurement Error’ assumption in the MR-Egger estimate; values above 0.9 indicate minimal bias, between 0.6 and 0.9 indicates some bias so we have run simulation extrapolation (SIMEX) corrections in place of MR-Egger where this is the case, and values under 0.6 indicate a larger bias where a SIMEX correction is also not appropriate.*

Table S2. Two-sample Mendelian randomisation results with smoking initiation as the exposure

| Outcome | Method | Number of SNPs | OR or beta^1^ (95% CI) | P-value | Heterogeneity test p-value^2^ | Directional pleiotropy intercept (95% CI; p-value) |
| --- | --- | --- | --- | --- | --- | --- |
| Drinks per week | IVW | 72 | 0.06  (0.03, 0.09) | 9.44x10^-06^ | 7.39x10^-15^ |  |
|  | MR Egger^a^ | - | - | - | - | - |
|  | Weighted median | 72 | 0.07  (0.05, 0.10) | 1.13x10^-07^ |  |  |
|  | Simple mode | 72 | 0.09  (0.02, 0.15) | 0.01 |  |  |
|  | Weighted mode | 72 | 0.09  (0.03, 0.14) | 0.002 |  |  |
| Cannabis use | IVW | 144 | 1.34  (1.24, 1.44) | 1.95x10^-14^ | 4.09x10^-08^ |  |
|  | MR Egger | 144 | 1.16  (0.97, 1.39) | 0.11 | 9.25x10^-08^ | 0.005  (-0.0008, 0.01; 0.09) |
|  | Weighted median | 144 | 1.14  (1.04, 1.26) | 0.005 |  |  |
|  | Simple mode | 144 | 1.20  (0.89, 1.60) | 0.22 |  |  |
|  | Weighted mode | 144 | 1.09  (0.96, 1.25) | 0.17 |  |  |
| Cannabis dependence | IVW | 133 | 1.68  (1.12, 2.51) | 0.01 | 0.56 |  |
|  | SIMEX adjusted MR Egger^b^ | 133 | 6.72  (0.95, 47.25) | 0.06 |  | -0.03  (-0.07, 0.01; 0.20) |
|  | Weighted median | 133 | 1.59  (0.88, 2.88) | 0.13 |  |  |
|  | Simple mode | 133 | 0.79  (0.16, 3.92) | 0.77 |  |  |
|  | Weighted mode | 133 | 0.94  (0.22, 4.00) | 0.93 |  |  |
| Cocaine dependence | IVW | 190 | 1.21  (0.58, 2.53) | 0.60 | 0.39 |  |
|  | SIMEX adjusted MR Egger^b^ | 190 | 28.61  (1.17, 699.55) | 0.04 |  | -0.06  (-0.13, 0.008; 0.08) |
|  | Weighted median | 190 | 0.89  (0.29, 2.73) | 0.84 |  |  |
|  | Simple mode | 190 | 0.33  (0.01, 10.90) | 0.54 |  |  |
|  | Weighted mode | 190 | 0.48  (0.02, 10.22) | 0.64 |  |  |
| Opioid dependence | IVW | 155 | 1.41  (0.62, 3.20) | 0.41 | 0.21 |  |
|  | SIMEX adjusted MR Egger^b^ | 155 | 16.72  (0.32, 886.86) | 0.17 |  | -0.05  (-0.13, 0.03; 0.24) |
|  | Weighted median | 155 | 1.89  (0.57, 6.21) | 0.30 |  |  |
|  | Simple mode | 155 | 0.87  (0.03, 25.19) | 0.94 |  |  |
|  | Weighted mode | 155 | 2.93  (0.23, 36.82) | 0.41 |  |  |

*^1^Beta for drinks per week, OR (odds ratio) for all other outcomes. ^2^For IVW this is Cochran’s test of heterogeneity and for MR-Egger this is Rucker’s Q-test. This is not presented where a SIMEX correction is reported. ^a^NO Measurement Error (NOME) assumption violated for MR-Egger and value below 0.6 so results are not presented. ^b^I-squared value between 0.6 and 0.9 so SIMEX correction is presented instead of MR-Egger.*

Table S3. Two-sample Mendelian randomisation results with smoking initiation as the outcome

| Exposure | Method | Number of SNPs | OR (95% CI) | P-value | Heterogeneity test p-value^1^ | Directional pleiotropy intercept (95% CI; p-value) |
| --- | --- | --- | --- | --- | --- | --- |
| Drinks per week | IVW | 22 | 1.26 (0.92, 1.72) | 0.15 | 2.09x10^-13^ |  |
|  | MR Egger | 22 | 1.07 (0.60, 1.93) | 0.82 | 2.17x10^-13^ | 0.002 (-1.03, 1.04; 0.53) |
|  | Weighted median | 22 | 0.98 (0.79, 1.23) | 0.88 |  |  |
|  | Simple mode | 22 | 0.84 (0.57, 1.24) | 0.35 |  |  |
|  | Weighted mode | 22 | 0.95 (0.77, 1.17) | 0.60 |  |  |
| Cannabis use | IVW | 4 | 1.39 (1.08, 1.80) | 0.01 | 2.56x10^-10^ |  |
|  | MR Egger^a^ | - | - | - | - | - |
|  | Weighted median | 4 | 1.21 (1.08, 1.35) | 8.13x10^-04^ |  |  |
|  | Simple mode | 4 | 1.17 (1.03, 1.32) | 0.07 |  |  |
|  | Weighted mode | 4 | 1.18 (1.05, 1.32) | 0.07 |  |  |
| Cannabis dependence | IVW | 9 | 1.00 (0.99, 1.01) | 0.60 | 0.04 |  |
|  | MR Egger^a^ | - | - | - | - | - |
|  | Weighted median | 9 | 1.00 (0.99, 1.01) | 0.86 |  |  |
|  | Simple mode | 9 | 1.00 (0.99, 1.01) | 0.68 |  |  |
|  | Weighted mode | 9 | 1.00 (0.99, 1.01) | 0.69 |  |  |
| Cocaine dependence | IVW | 27 | 1.00 (1.00, 1.00) | 0.42 | 0.42 |  |
|  | SIMEX adjusted MR Egger^b^ | 27 | 1.00 (1.00, 1.00) | 0.76 |  | 0.0005 (-0.004, 0.005; 0.82) |
|  | Weighted median | 27 | 1.00 (1.00, 1.00) | 0.29 |  |  |
|  | Simple mode | 27 | 1.00 (1.00, 1.00) | 0.48 |  |  |
|  | Weighted mode | 27 | 1.00 (1.00, 1.00) | 0.49 |  |  |
| Opioid dependence | IVW | 7 | 1.00 (0.99, 1.01) | 0.80 | 0.0005 |  |
|  | MR Egger^a^ | - | - | - | - | - |
|  | Weighted median | 7 | 1.00 (0.99, 1.00) | 0.20 |  |  |
|  | Simple mode | 7 | 1.01 (0.99, 1.02) | 0.38 |  |  |
|  | Weighted mode | 7 | 1.00 (0.99, 1.00) | 0.28 |  |  |

*^1^For IVW this is Cochran’s test of heterogeneity and for MR-Egger this is Rucker’s Q-test. This is not presented where a SIMEX correction is reported. ^a^NO Measurement Error (NOME) assumption violated for MR-Egger and value below 0.6 so results are not presented. ^b^I-squared value between 0.6 and 0.9 so SIMEX correction is presented instead of MR-Egger.*

Table S4. Two-sample Mendelian randomisation results with drinks per week as the exposure

| Outcome | Method | Number of SNPs | OR (95% CI) | P-value | Heterogeneity test p-value^1^ | Directional pleiotropy intercept (95% CI; p-value) |
| --- | --- | --- | --- | --- | --- | --- |
| Cannabis use | IVW | 20 | 0.55 (0.16, 1.93) | 0.35 | 3.40x10-06 |  |
|  | MR Egger | 20 | 0.08 (0.01 0.48) | 0.01 | 7.94x10-04 | 0.02 (0.005, 0.03; 0.01) |
|  | Weighted median | 20 | 0.24 (0.09, 0.64) | 0.004 |  |  |
|  | Simple mode | 20 | 0.45 (0.03, 5.96) | 0.55 |  |  |
|  | Weighted mode | 20 | 0.18 (0.07, 0.47) | 0.002 |  |  |
| Cannabis dependence | IVW | 41 | 2.73 (0.62, 11.95) | 0.18 | 0.26 |  |
|  | SIMEX adjusted MR Egger^b^ | 41 | 12.46 (0.14, 1088.49) | 0.27 |  | -0.02 (-0.07, 0.03; 0.50) |
|  | Weighted median | 41 | 3.96 (0.52, 30.38) | 0.19 |  |  |
|  | Simple mode | 41 | 2.70 (0.05, 134.58) | 0.62 |  |  |
|  | Weighted mode | 41 | 3.44 (0.19, 61.45) | 0.41 |  |  |
| Cocaine dependence | IVW | 68 | 0.50 (0.09, 2.79) | 0.43 | 0.15 |  |
|  | MR Egger | 68 | 0.62 (0.05, 8.25) | 0.72 | 0.13 | -0.005 (-0.05, 0.04; 0.82) |
|  | Weighted median | 68 | 0.52 (0.05, 5.04) | 0.57 |  |  |
|  | Simple mode | 68 | 105.78 (0.03, 3.68x10^05^) | 0.27 |  |  |
|  | Weighted mode | 68 | 0.53 (0.05, 5.37) | 0.59 |  |  |
| Opioid dependence | IVW | 47 | 0.38 (0.06, 2.41) | 0.30 | 0.74 |  |
|  | MR Egger | 47 | 0.11 (0.007, 1.48) | 0.10 | 0.77 | 0.03 (-0.01, 0.07; 0.19) |
|  | Weighted median | 47 | 0.12 (0.01, 1.24) | 0.07 |  |  |
|  | Simple mode | 47 | 0.15 (0.0001, 171.96) | 0.60 |  |  |
|  | Weighted mode | 47 | 0.10 (0.009, 1.22) | 0.08 |  |  |

*^1^For IVW this is Cochran’s test of heterogeneity and for MR-Egger this is Rucker’s Q-test. This is not presented where a SIMEX correction is reported. ^a^NO Measurement Error (NOME) assumption violated for MR-Egger and value below 0.6 so results are not presented. ^b^I-squared value between 0.6 and 0.9 so SIMEX correction is presented instead of MR-Egger.*

Table S5. Two-sample Mendelian randomisation results with drinks per week as the outcome

| Exposure | Method | Number of SNPs | Beta (95% CI) | P-value | Heterogeneity test p-value^1^ | Directional pleiotropy intercept (95% CI; p-value) |
| --- | --- | --- | --- | --- | --- | --- |
| Cannabis use | IVW | 4 | 0.03 (-0.009, 0.07) | 0.14 | 2.43 x10-06 |  |
|  | MR Egger^a^ | - | - | - | - | - |
|  | Weighted median | 4 | 0.03 (0.008, 0.05) | 0.006 |  |  |
|  | Simple mode | 4 | 0.03 (0.001, 0.06) | 0.14 |  |  |
|  | Weighted mode | 4 | 0.03 (0.005, 0.06) | 0.11 |  |  |
| Cannabis dependence | IVW | 9 | -0.0003 (-0.003, 0.002) | 0.80 | 0.22 |  |
|  | MR Egger^a^ | - | - | - | - | - |
|  | Weighted median | 9 | -0.001 (-0.004, 0.002) | 0.37 |  |  |
|  | Simple mode | 9 | -0.002 (-0.006, 0.003) | 0.43 |  |  |
|  | Weighted mode | 9 | -0.002 (-0.007, 0.003) | 0.44 |  |  |
| Cocaine dependence | IVW | 27 | 0.0007 (-0.00007, 0.001) | 0.08 | 0.11 |  |
|  | SIMEX adjusted MR Egger^b^ | 27 | -0.0001 (-0.002, 0.002) | 0.92 |  | 0.0009 (-0.001, 0.003; 0.46) |
|  | Weighted median | 27 | 0.0005 (-0.0004, 0.001) | 0.29 |  |  |
|  | Simple mode | 27 | 0.0009 (-0.0005, 0.001) | 0.36 |  |  |
|  | Weighted mode | 27 | 0.0007 (-0.001, 0.003) | 0.39 |  |  |
| Opioid dependence | IVW | 7 | 0.002 (0.0005, 0.003) | 8.61x10^-03^ | 0.33 |  |
|  | MR Egger^a^ | - | - | - | - | - |
|  | Weighted median | 7 | 0.002 (-0.0002, 0.004) | 0.08 |  |  |
|  | Simple mode | 7 | 0.002 (-0.0008, 0.006) | 0.20 |  |  |
|  | Weighted mode | 7 | 0.003 (-0.0005, 0.006) | 0.14 |  |  |

*^1^For IVW this is Cochran’s test of heterogeneity and for MR-Egger this is Rucker’s Q-test. This is not presented where a SIMEX correction is reported. ^a^NO Measurement Error (NOME) assumption violated for MR-Egger and value below 0.6 so results are not presented. ^b^I-squared value between 0.6 and 0.9 so SIMEX correction is presented instead of MR-Egger.*

Figure S1. Mendelian randomisation results with smoking initiation as the exposure and drinks per week as the outcome.

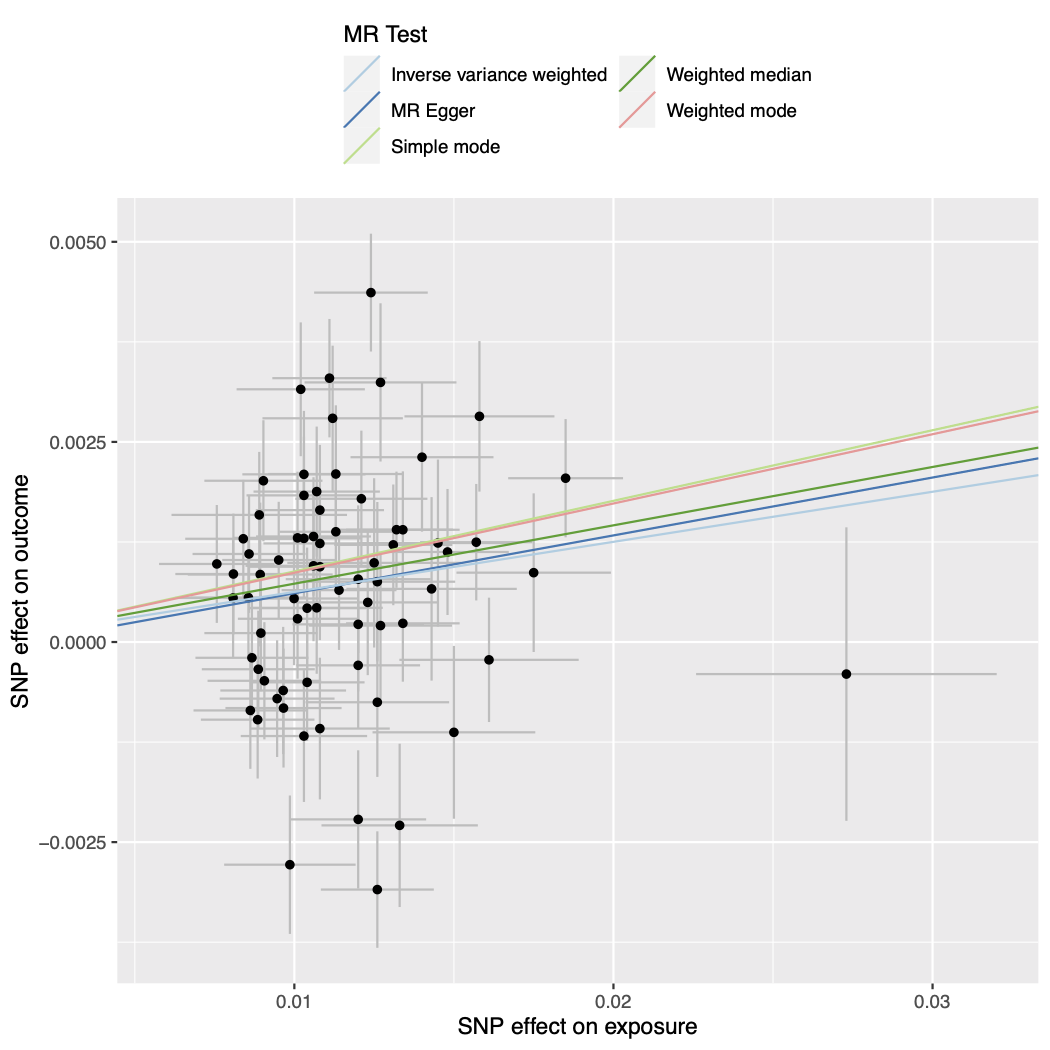

Figure S2. Forest plot of single SNP analysis using the Wald ratio with smoking initiation as the exposure and drinks per week as the outcome.

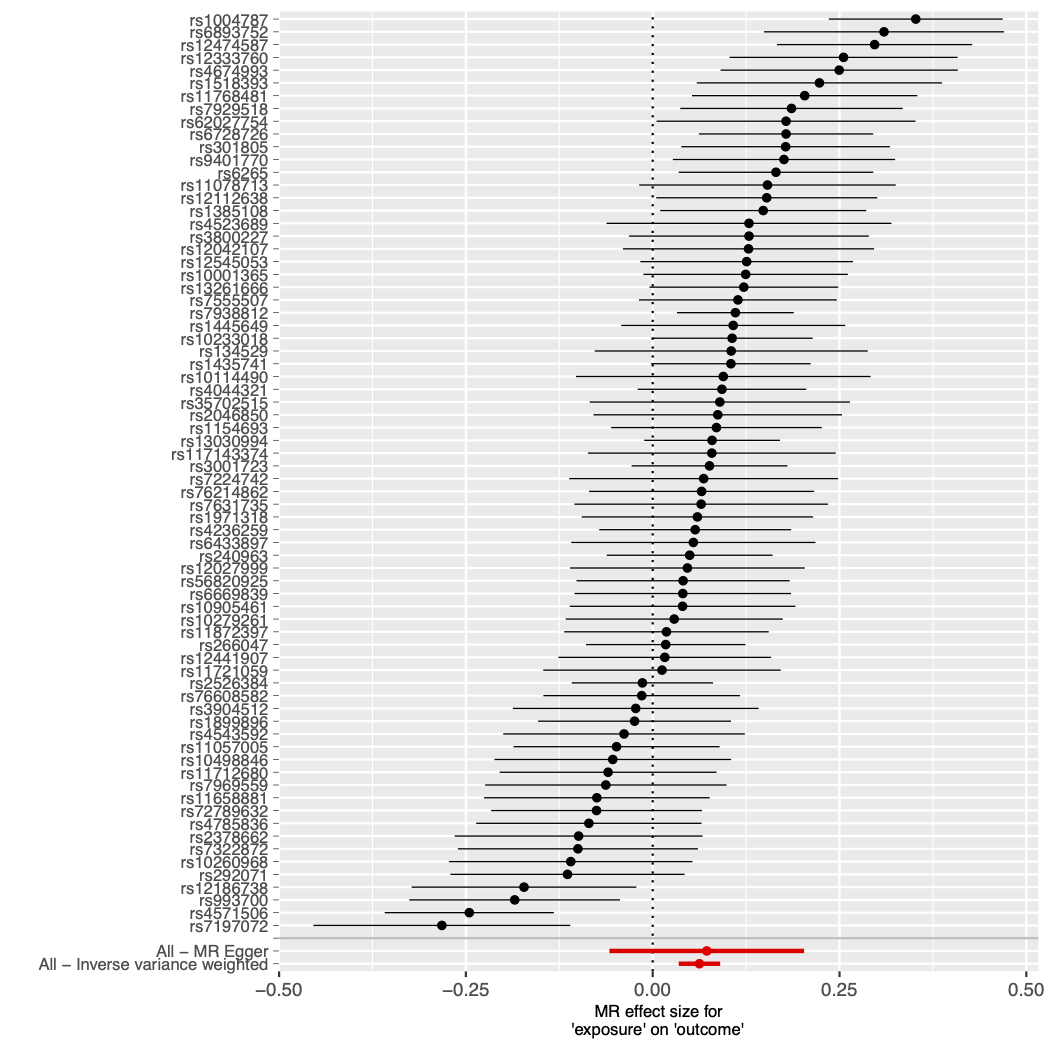

Figure S3. Funnel plot of individual Wald ratios for each SNP against their precision with smoking initiation as the exposure and drinks per week as the outcome.

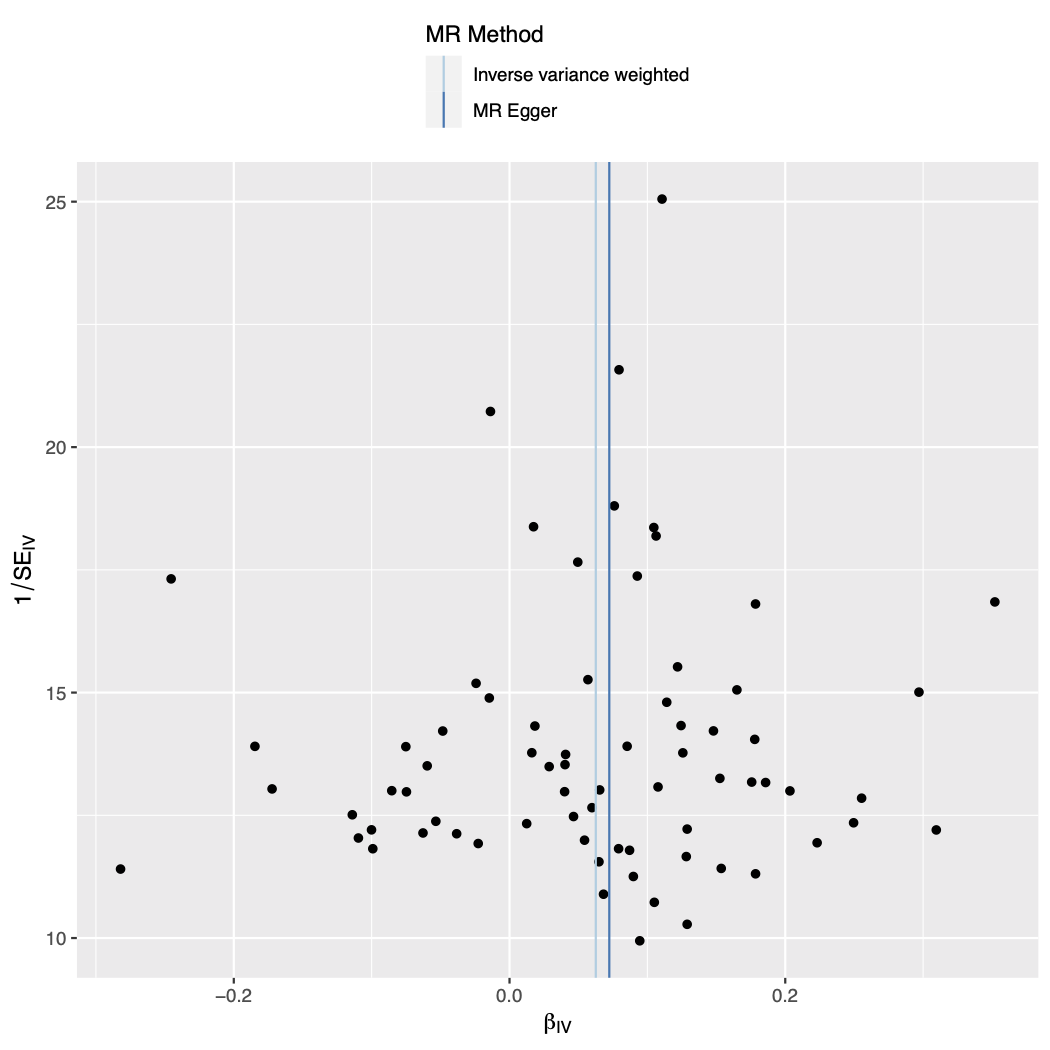

Figure S4. Leave-one-out analysis plot with smoking initiation as the exposure and drinks per week as the outcome.

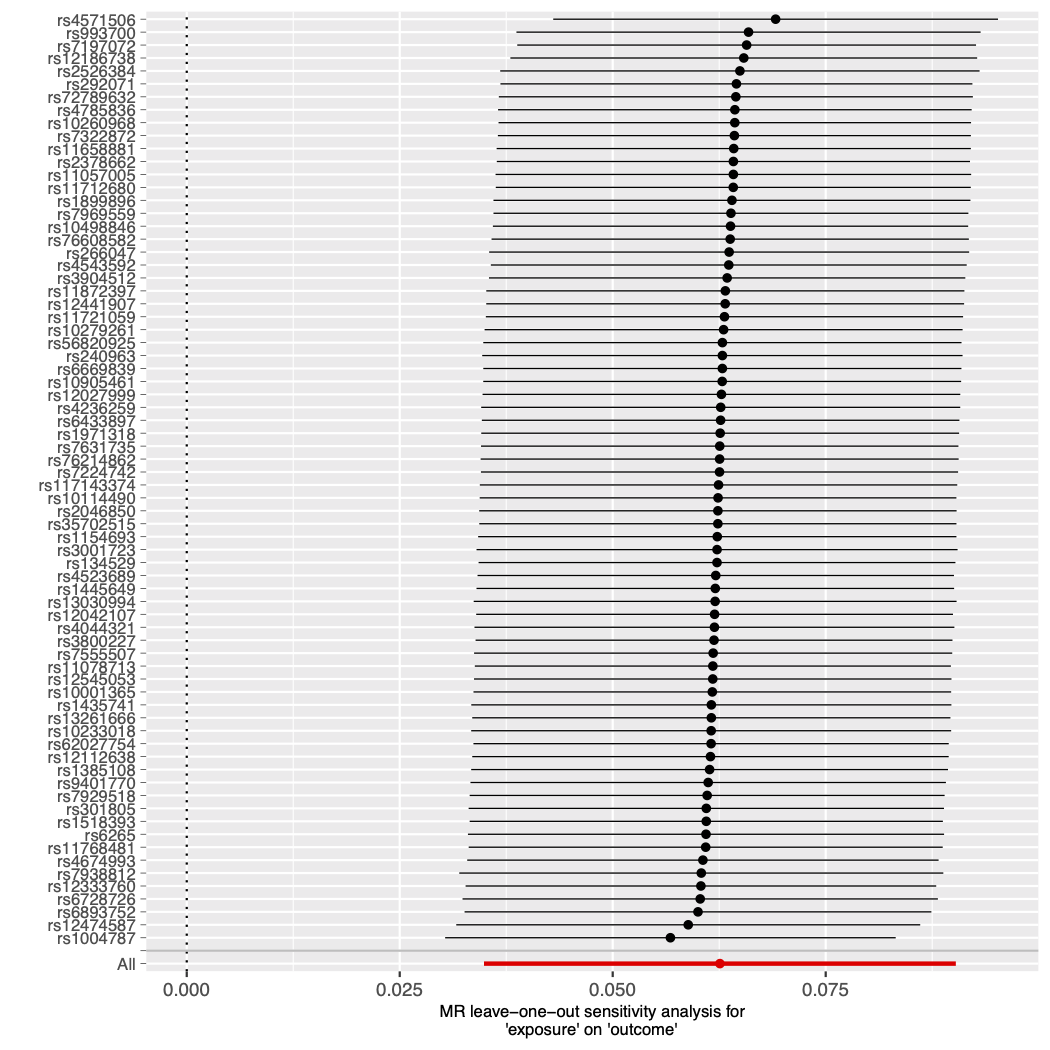

Figure S5. Mendelian randomisation results with smoking initiation as the exposure and cannabis use as the outcome.

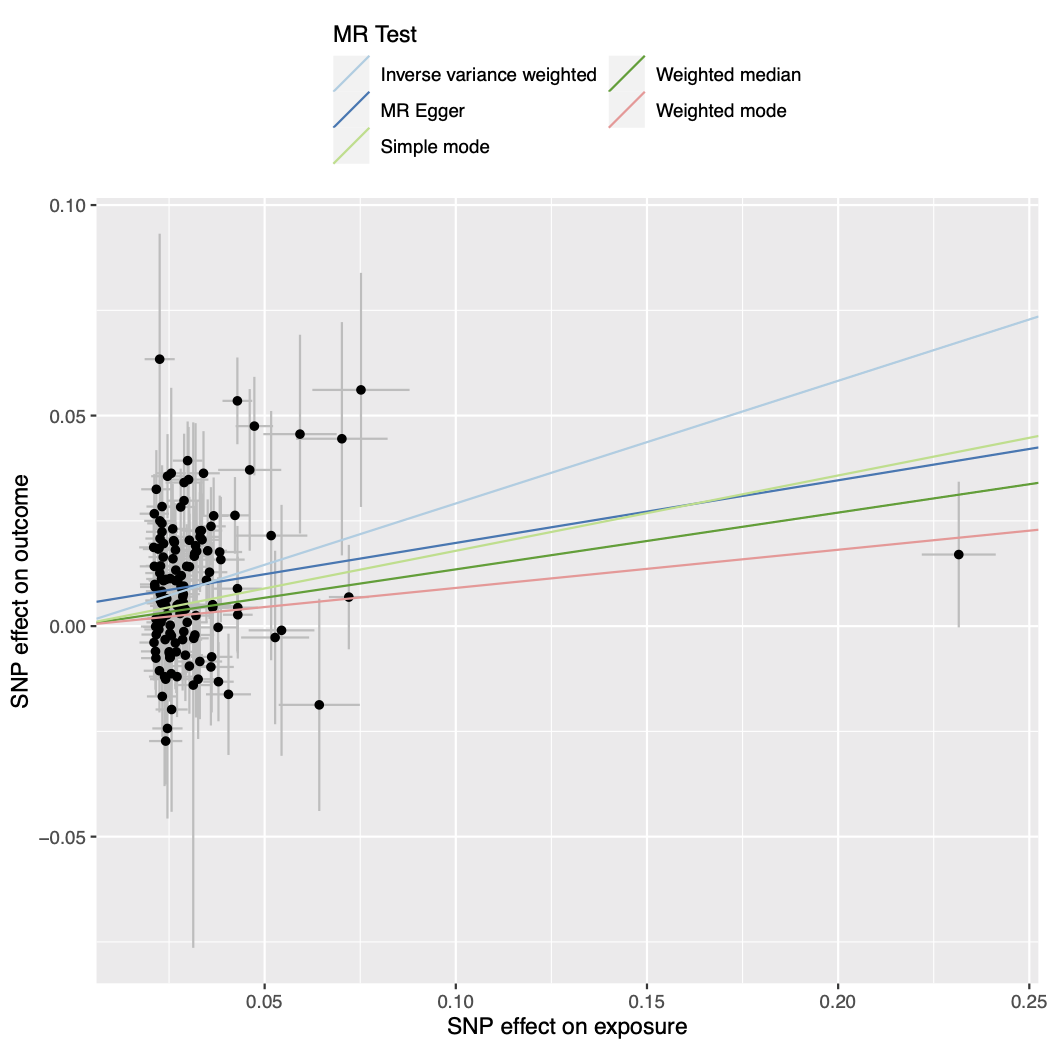

Figure S6. Forest plot of single SNP analysis using the Wald ratio with smoking initiation as the exposure and cannabis use as the outcome.

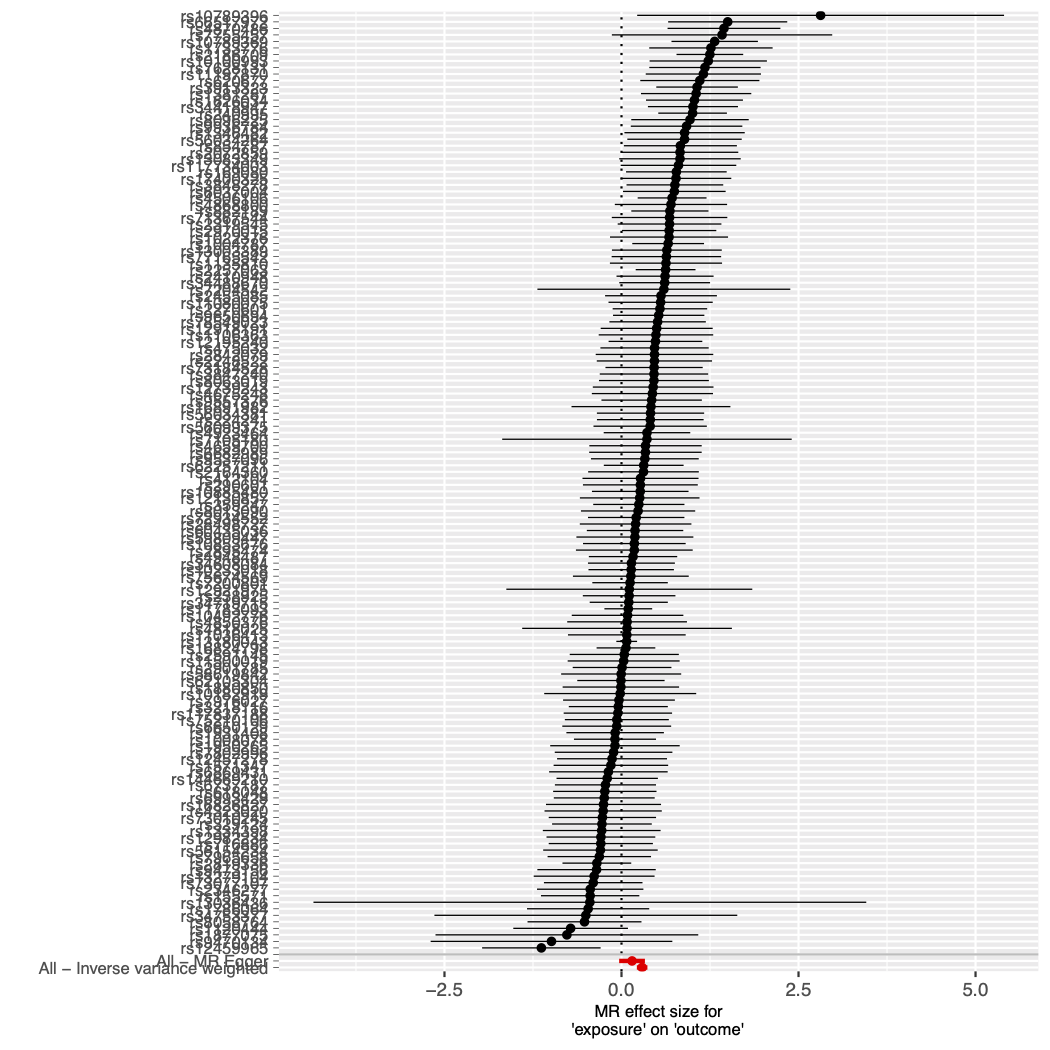

Figure S7. Funnel plot of individual Wald ratios for each SNP against their precision with smoking initiation as the exposure and cannabis use as the outcome.

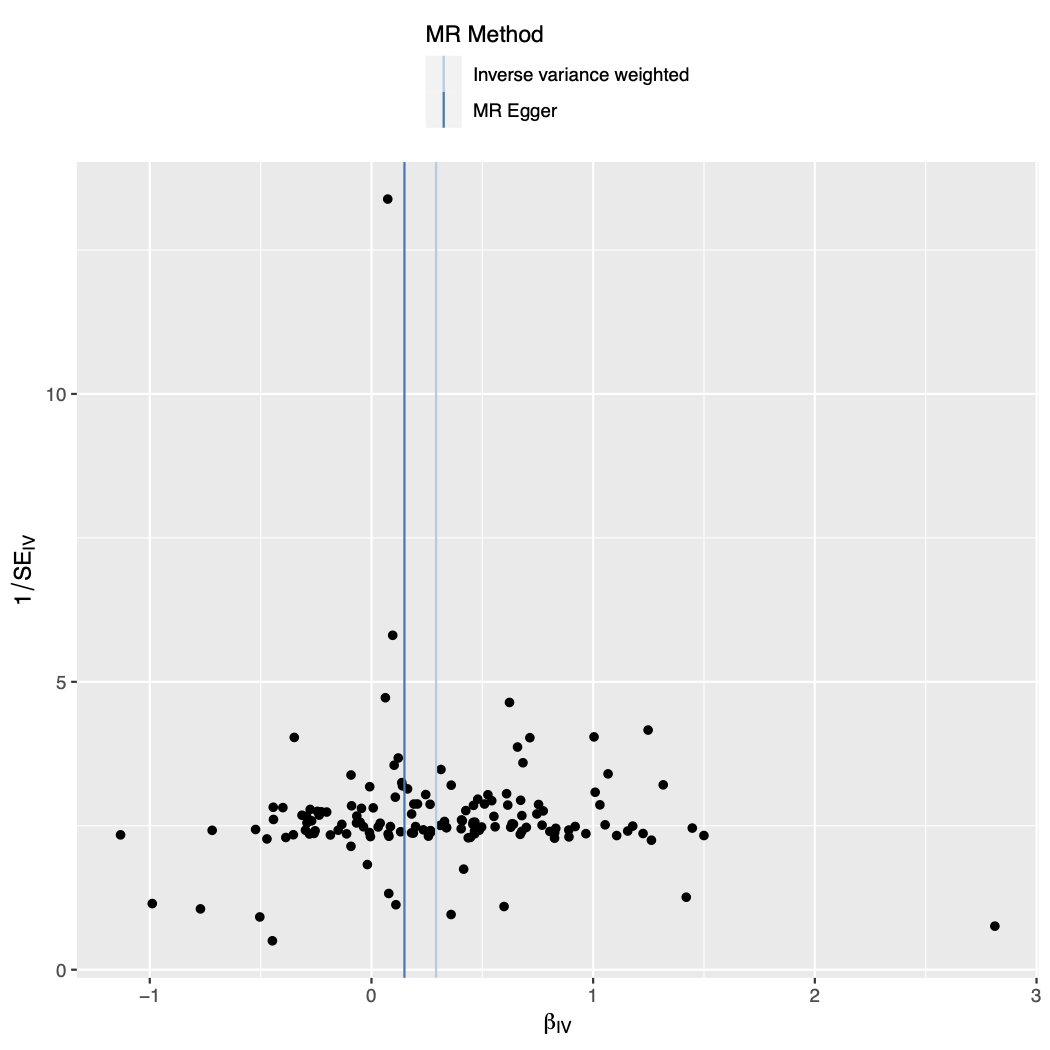

Figure S8. Leave-one-out analysis plot with smoking initiation as the exposure and cannabis use as the outcome.

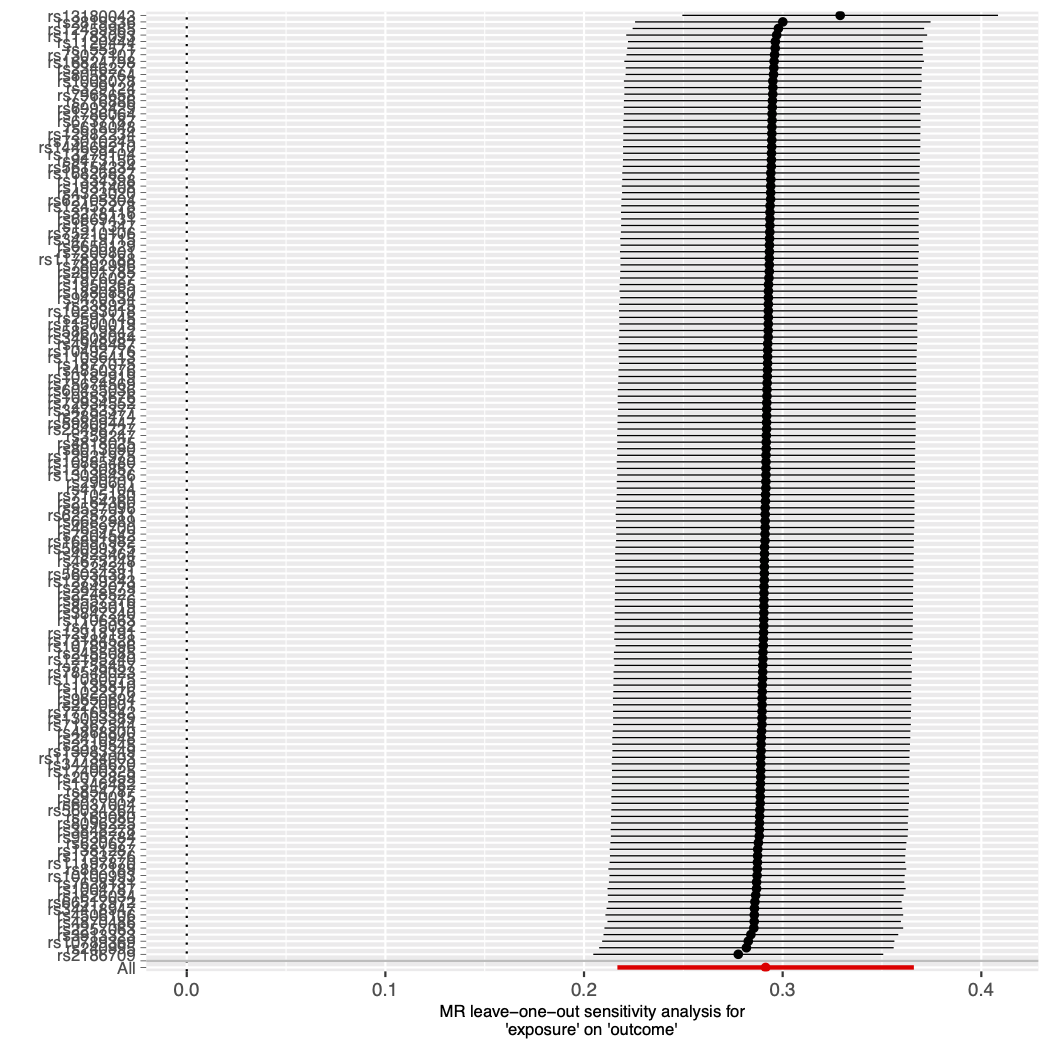

Figure S9. Mendelian randomisation results with smoking initiation as the exposure and cannabis dependence as the outcome.

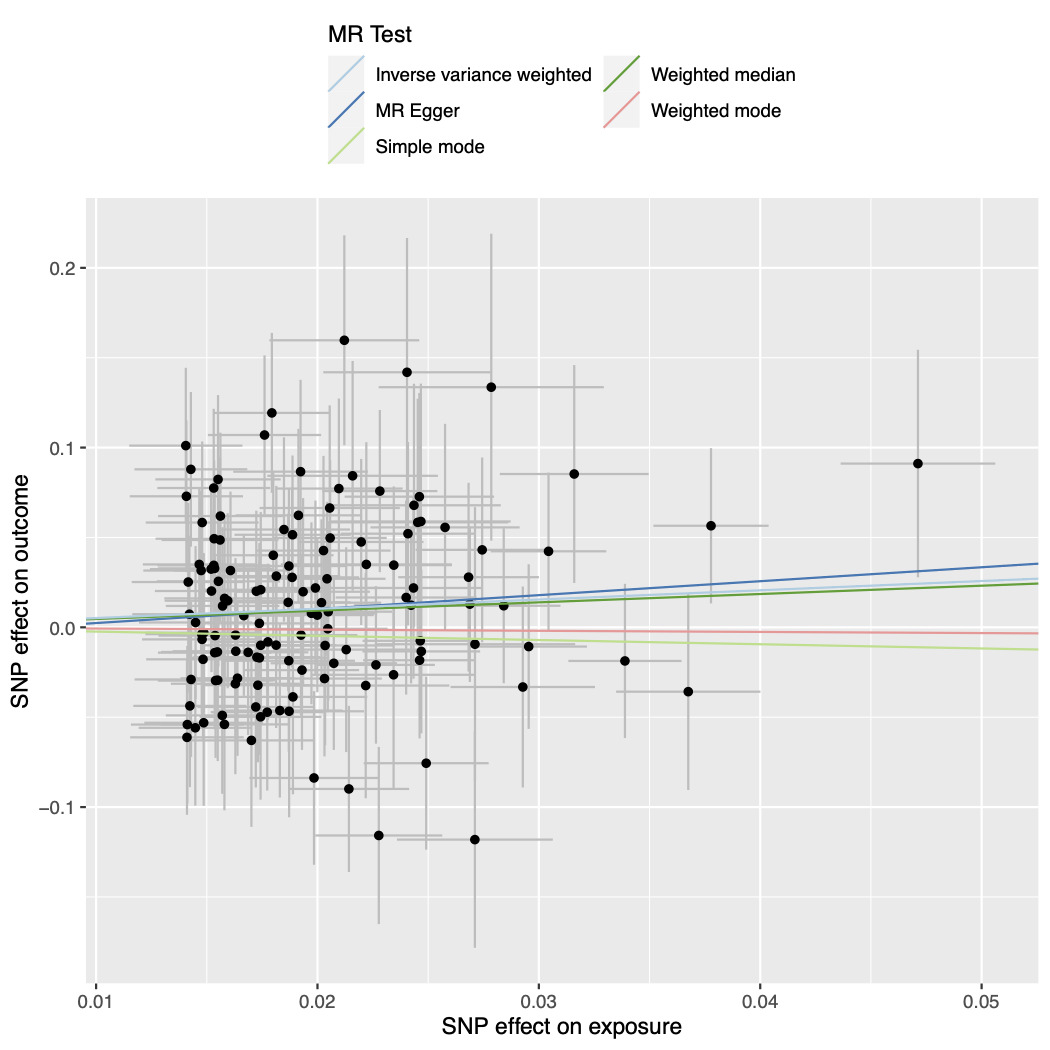

Figure S10. Forest plot of single SNP analysis using the Wald ratio with smoking initiation as the exposure and cannabis dependence as the outcome.

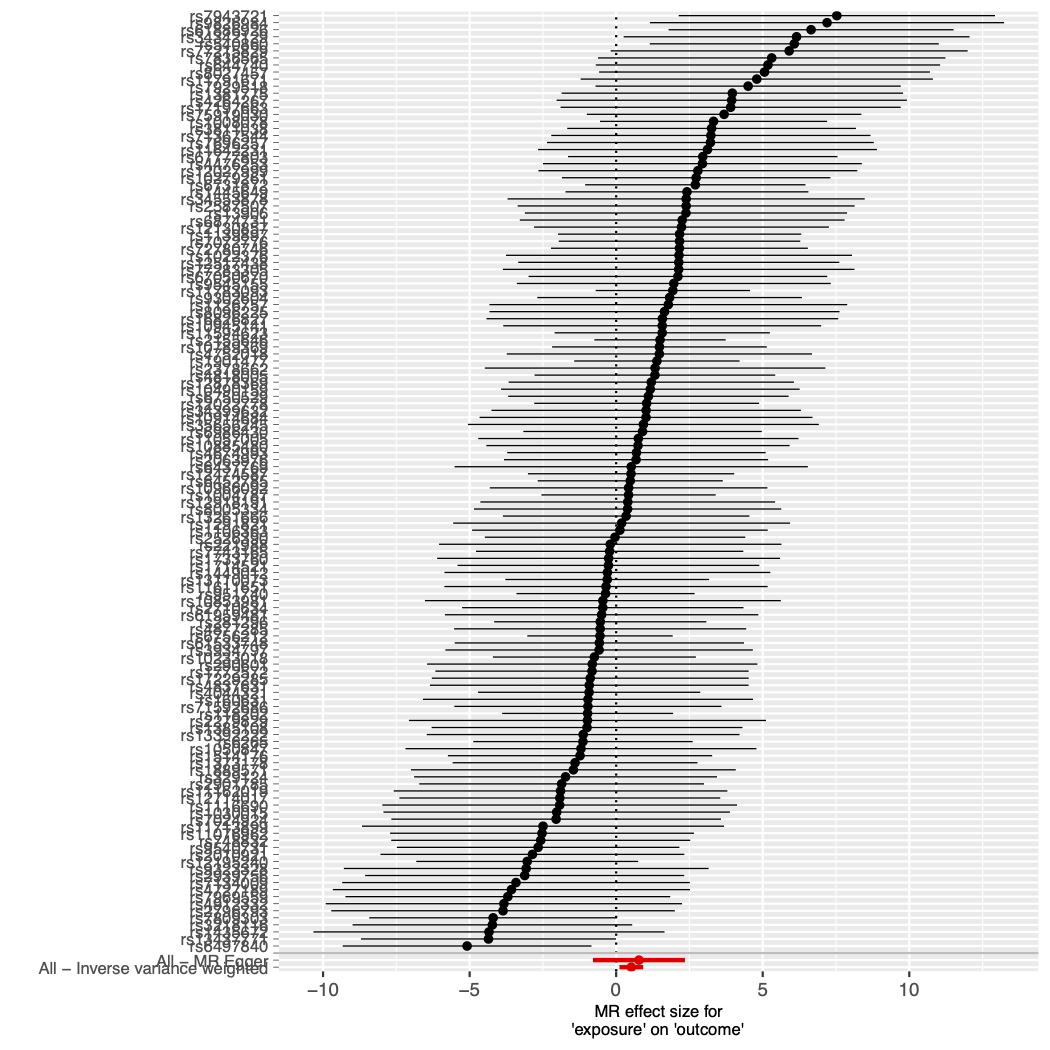

Figure S11. Funnel plot of individual Wald ratios for each SNP against their precision with smoking initiation as the exposure and cannabis dependence as the outcome.

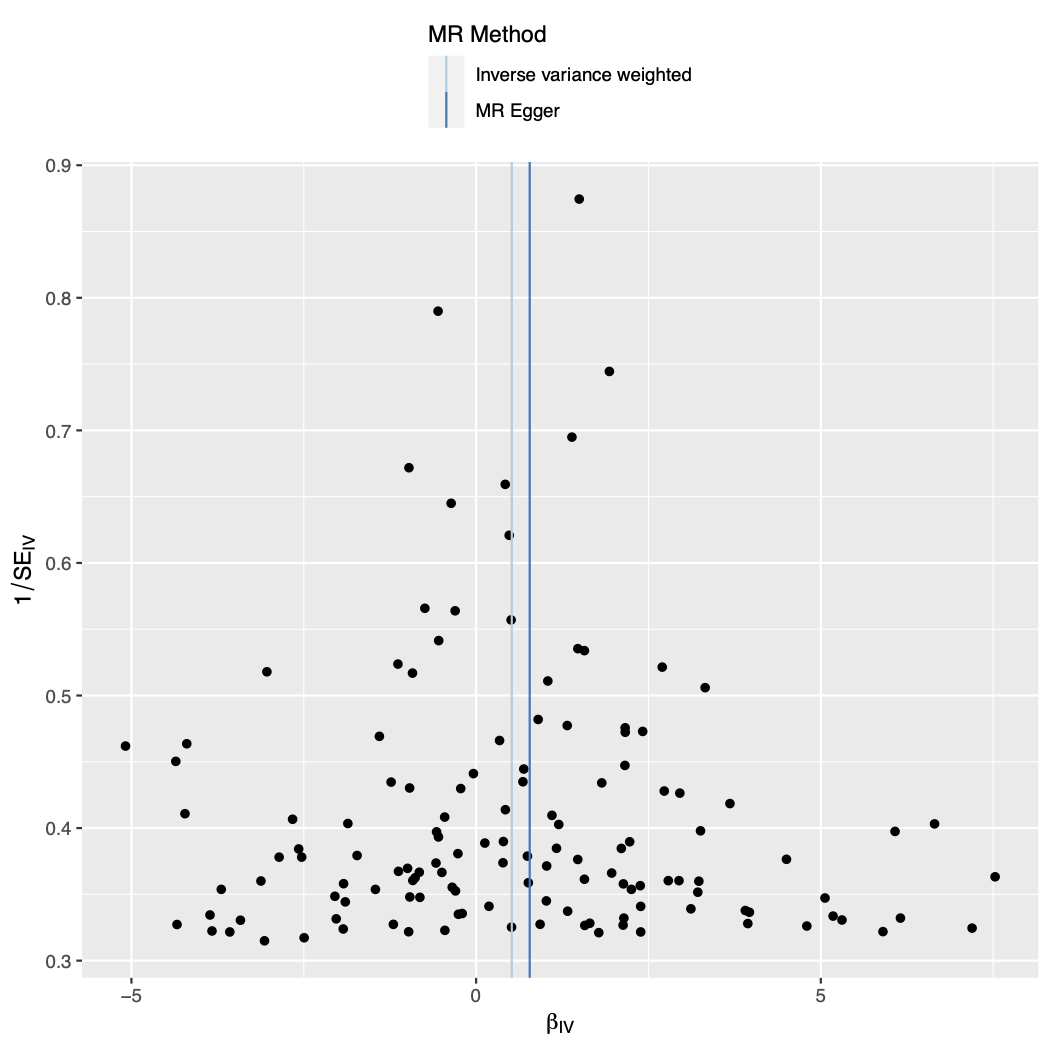

Figure S12. Leave-one-out analysis plot with smoking initiation as the exposure and cannabis dependence as the outcome.

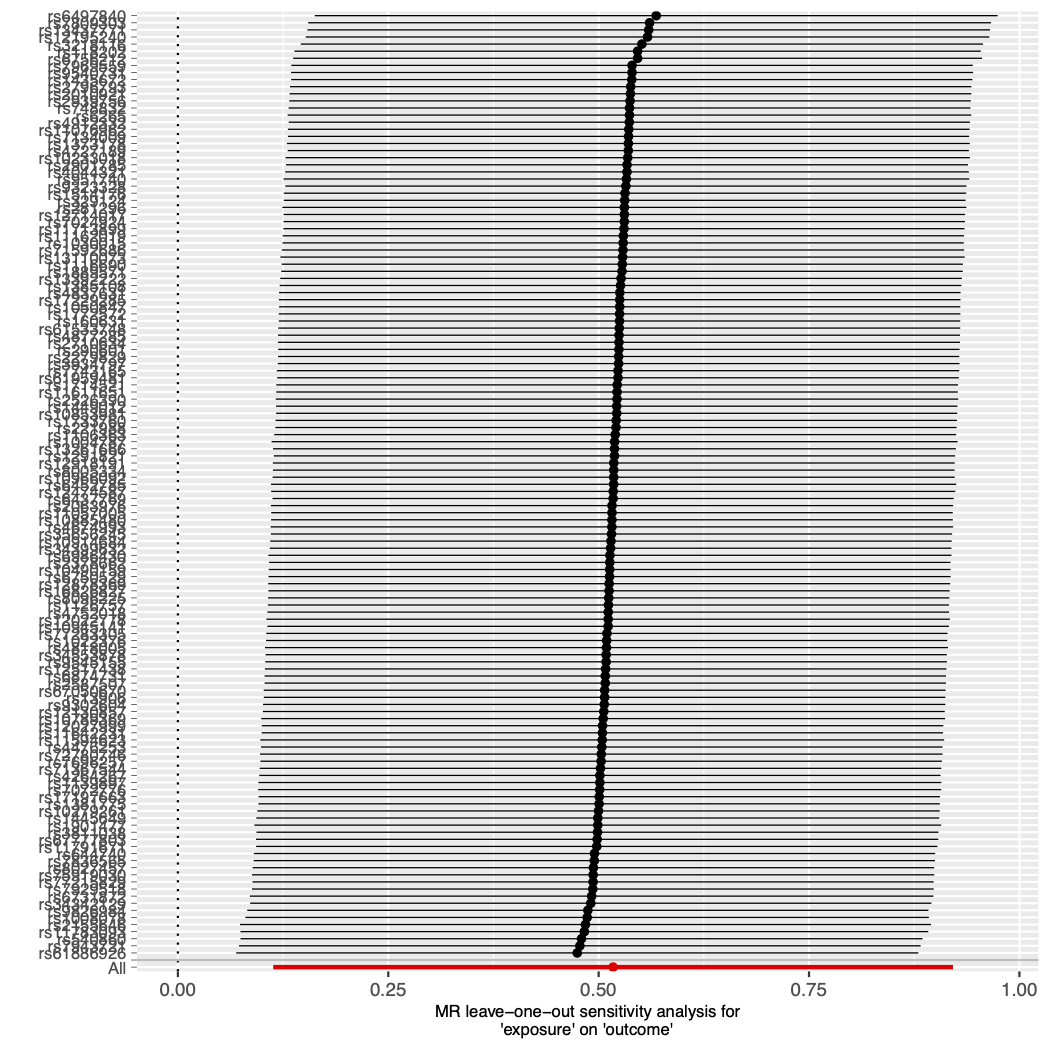

Figure S13. Mendelian randomisation results with cannabis use as the exposure and smoking initiation as the outcome.

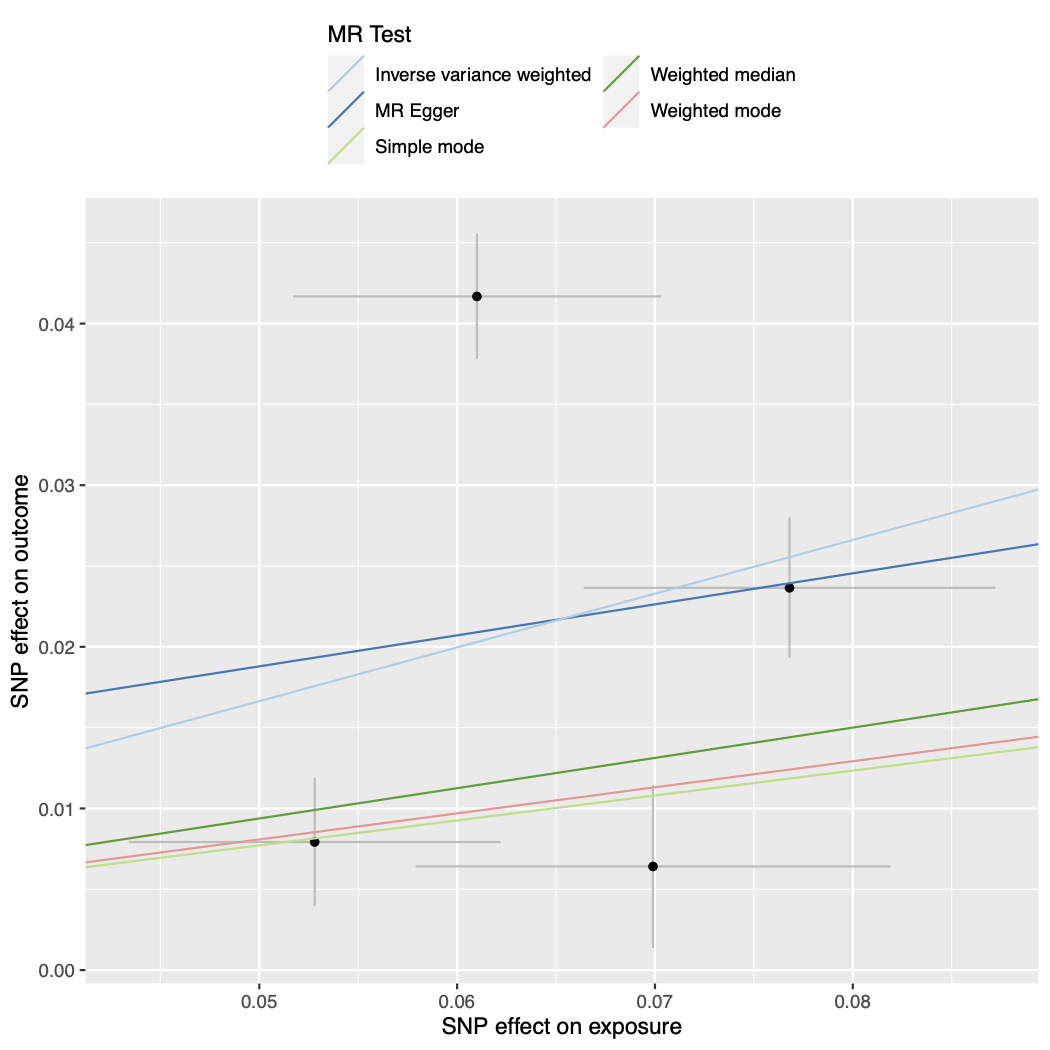

Figure S14. Funnel plot of individual Wald ratios for each SNP against their precision with cannabis use as the exposure and smoking initiation as the outcome.

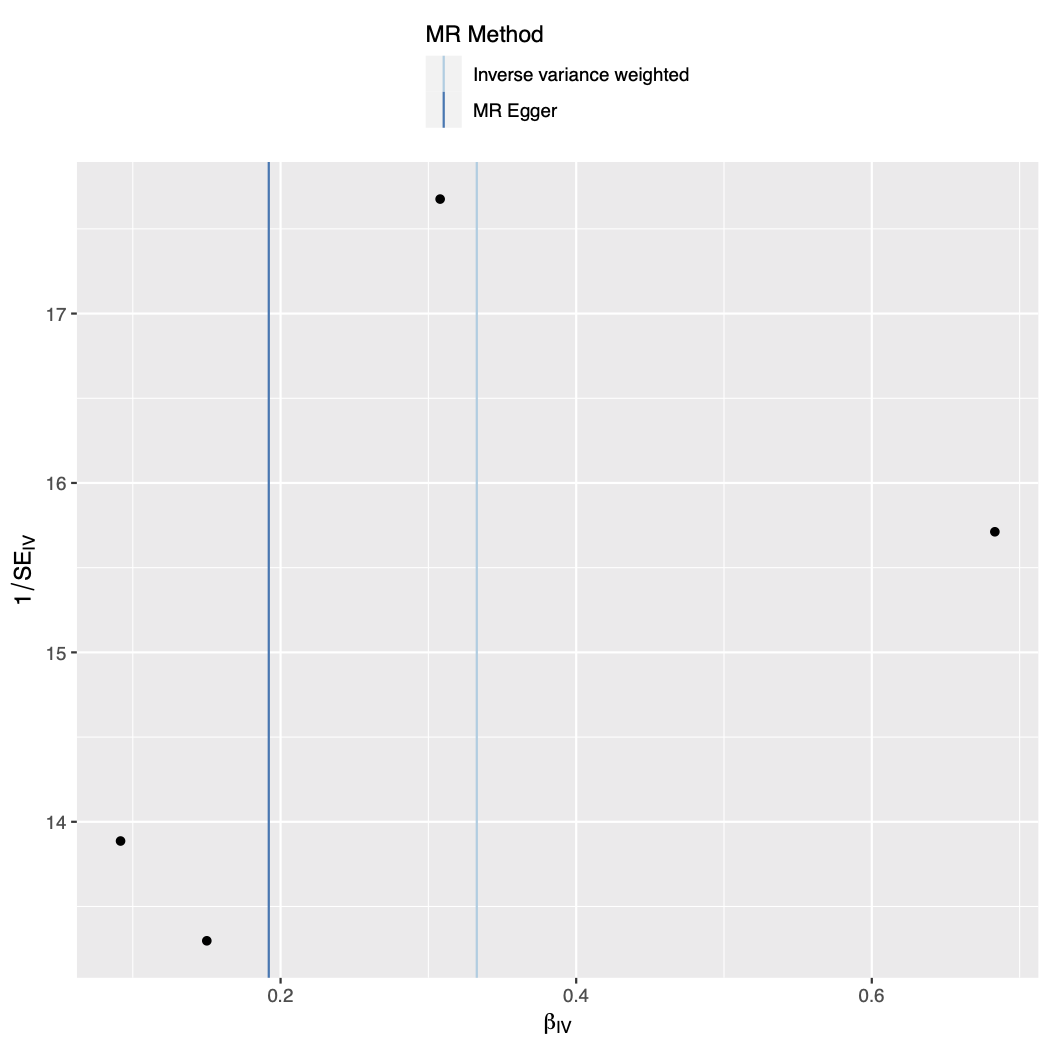

Figure S15. Forest plot of single SNP analysis using the Wald ratio with cannabis use as the exposure and smoking initiation as the outcome.

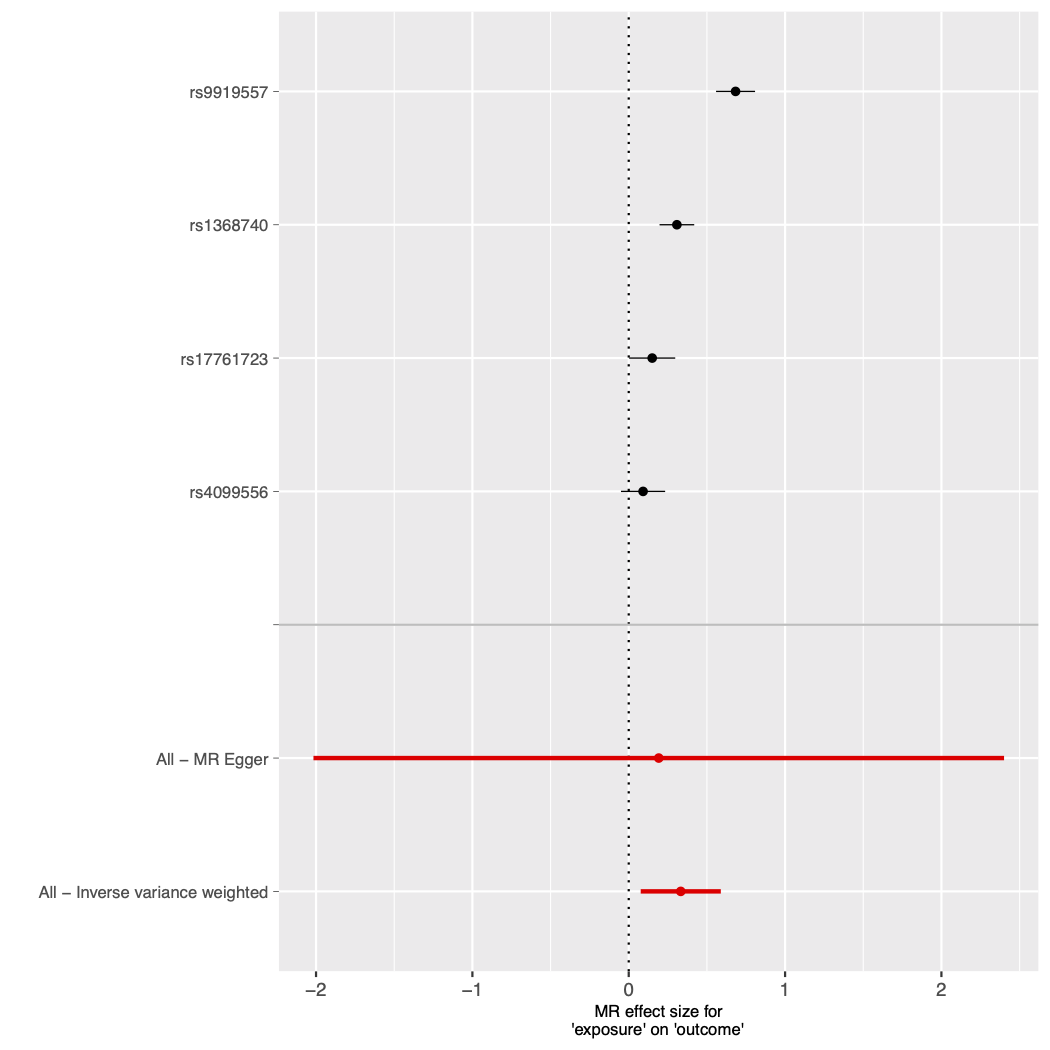

Figure S16. Leave-one-out analysis plot with cannabis use as the exposure and smoking initiation as the outcome.

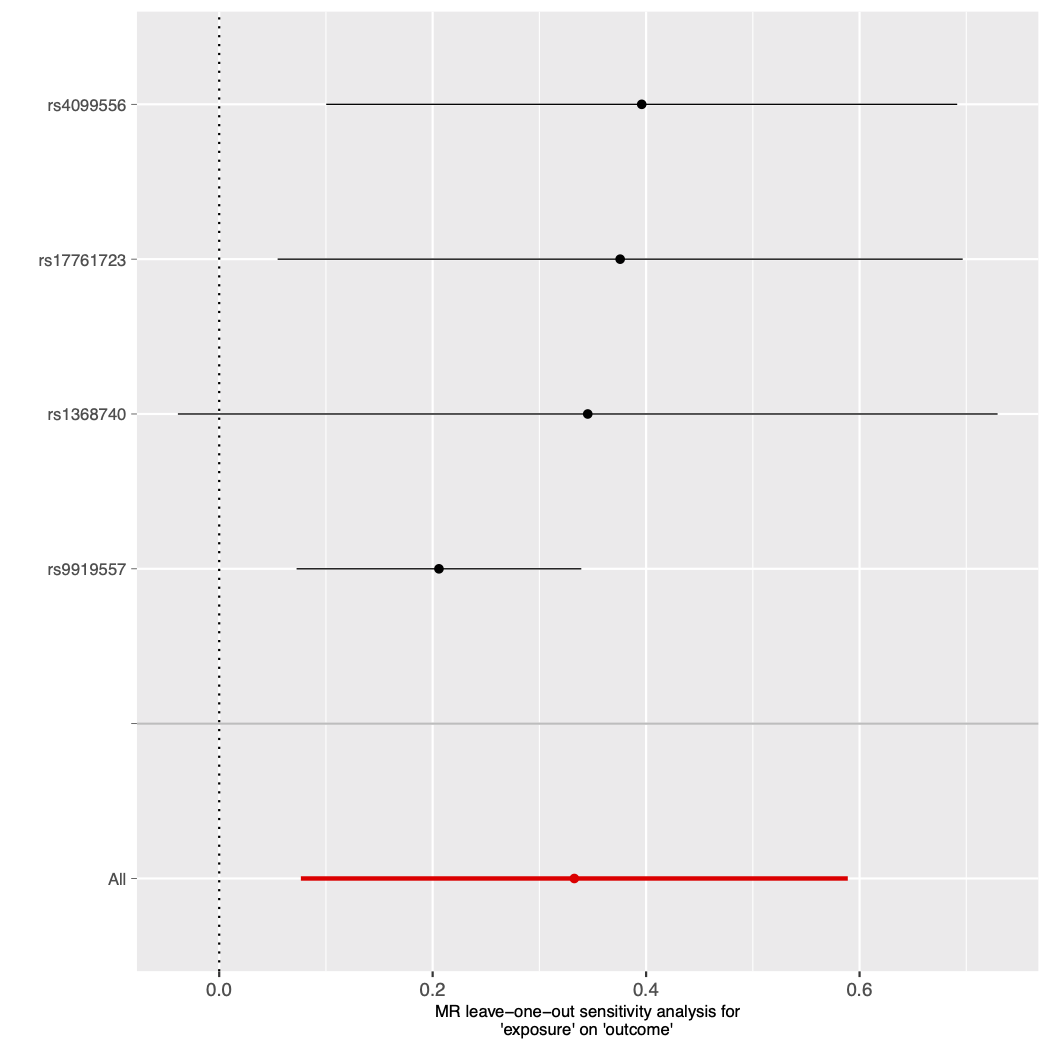

Figure S17. Mendelian randomisation results with opioid dependence as the exposure and drinks per week as the outcome.

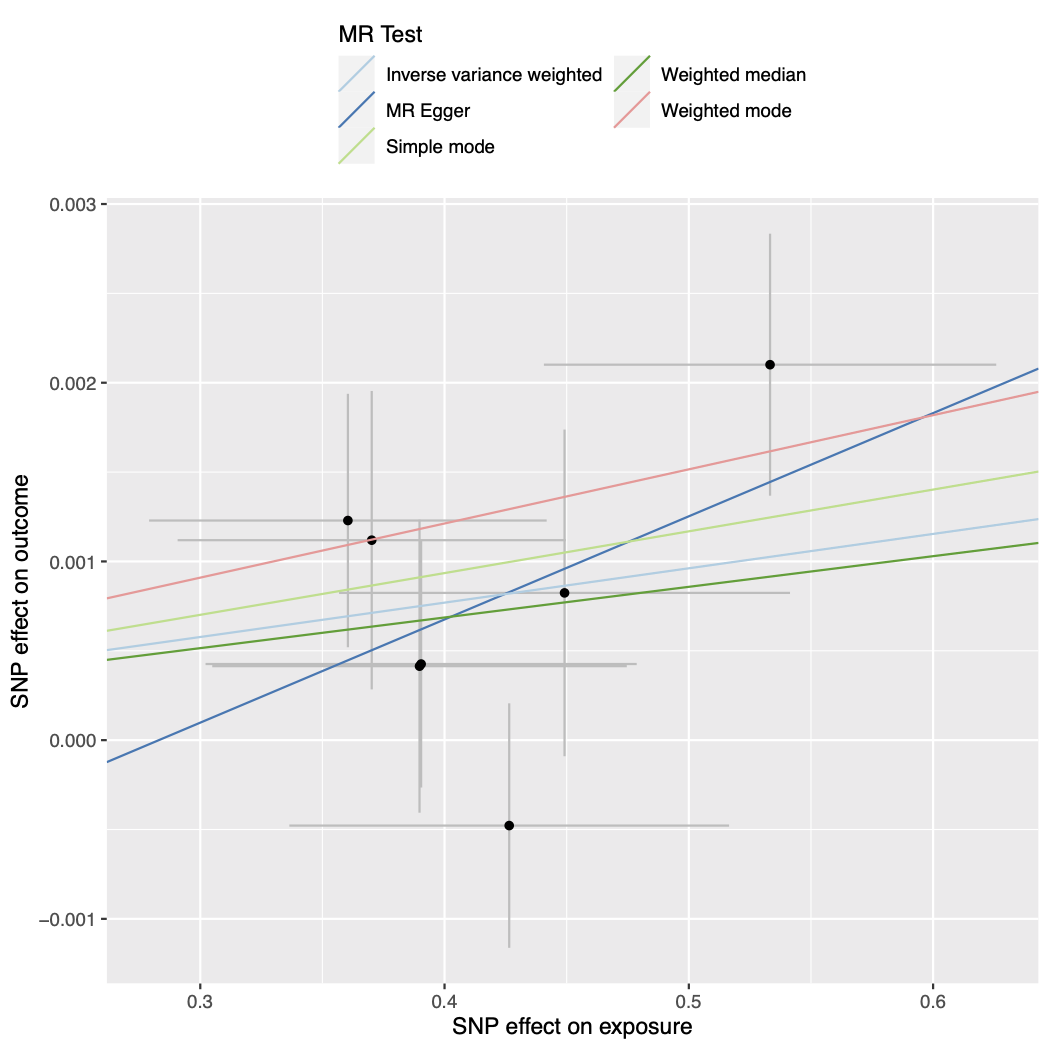

Figure S18. Forest plot of single SNP analysis using the Wald ratio with opioid dependence as the exposure and drinks per week as the outcome.

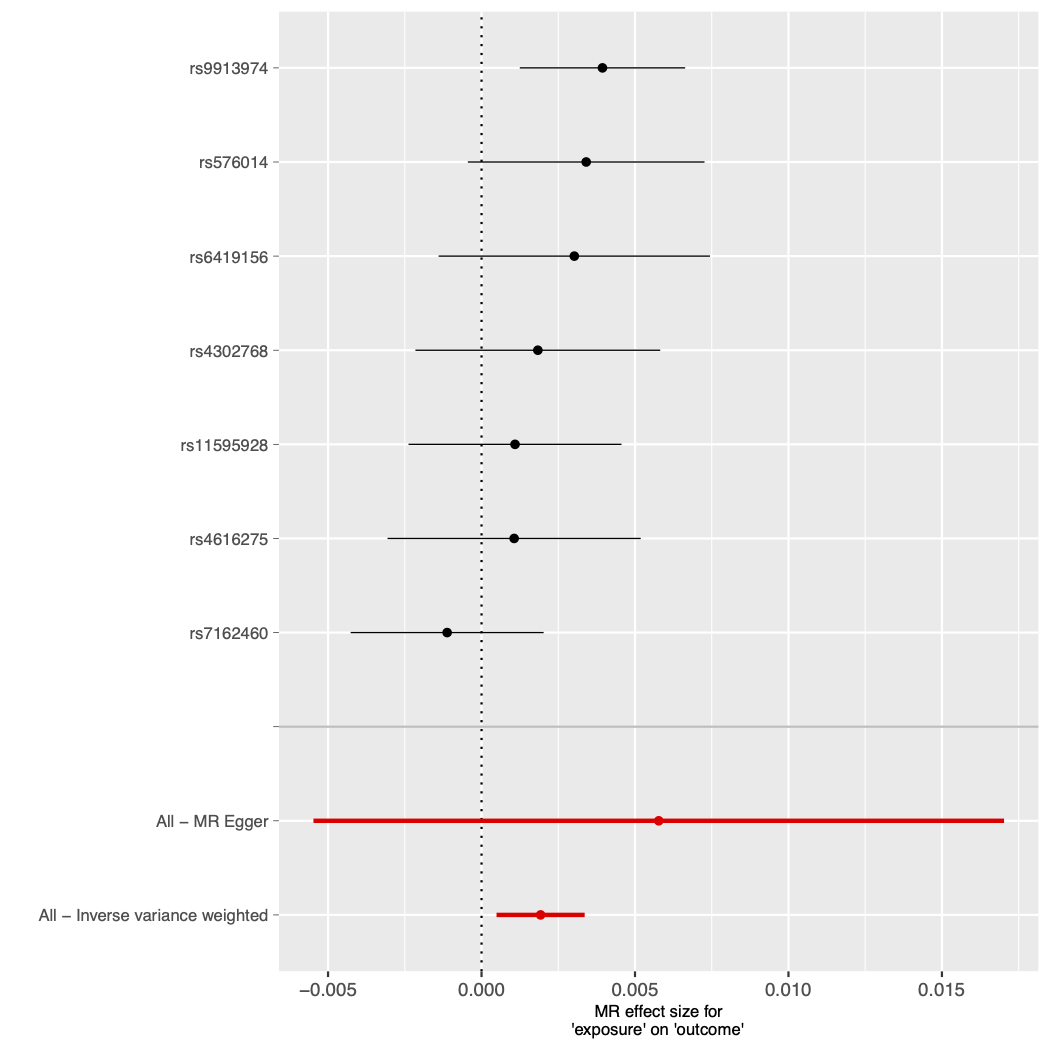

Figure S19. Funnel plot of individual Wald ratios for each SNP against their precision with opioid dependence as the exposure and drinks per week as the outcome.

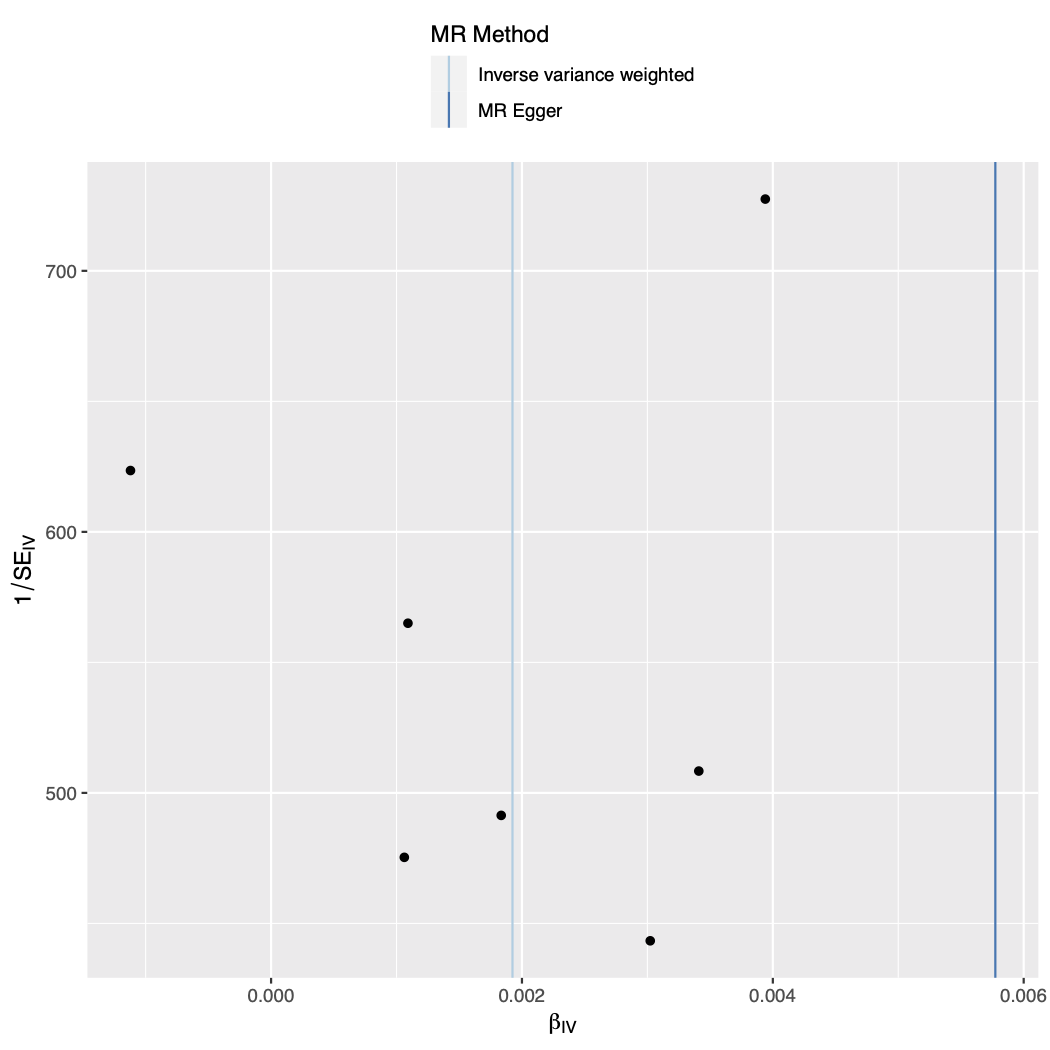

Figure S20. Leave-one-out analysis plot with opioid dependence as the exposure and drinks per week as the outcome.

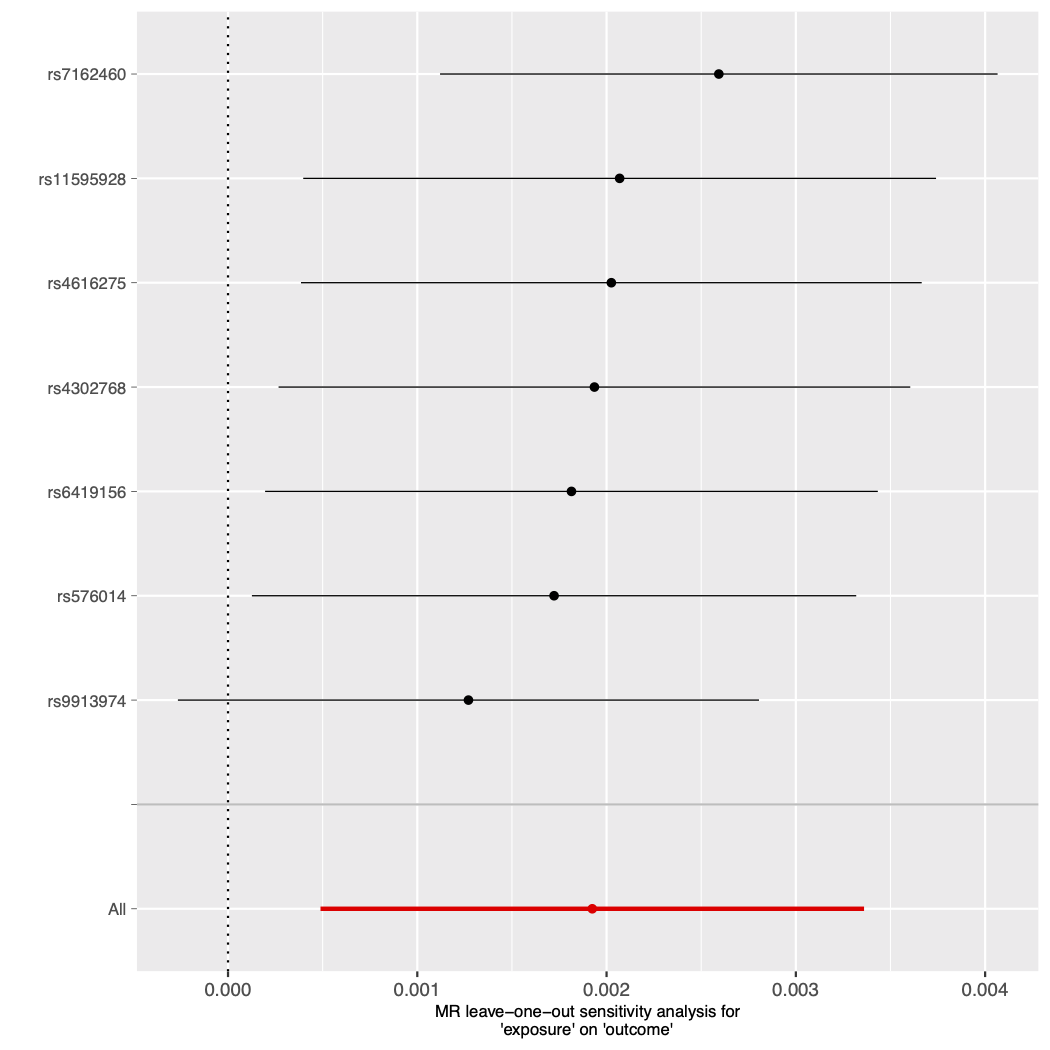
